# Safety of Monovalent BNT162b2 (Pfizer-BioNTech), mRNA-1273 (Moderna), and NVX-CoV2373 (Novavax) COVID-19 Vaccines in US Children Aged 6 months to 17 years

**DOI:** 10.1101/2023.10.13.23296903

**Authors:** Mao Hu, Azadeh Shoaibi, Yuhui Feng, Patricia C. Lloyd, Hui Lee Wong, Elizabeth R. Smith, Kandace L. Amend, Annemarie Kline, Daniel C. Beachler, Joann F. Gruber, Mahasweta Mitra, John D. Seeger, Charlalynn Harris, Alex Secora, Joyce Obidi, Jing Wang, Jennifer Song, Cheryl N. McMahill-Walraven, Christian Reich, Rowan McEvoy, Rose Do, Yoganand Chillarige, Robin Clifford, Danielle D Cooper, Richard Forshee, Steven A. Anderson

**Affiliations:** Acumen LLC, Burlingame, CA, USA; US Food and Drug Administration, Silver Spring, MD, USA; Optum Epidemiology, Boston, MA, USA; Carelon Research, Wilmington, DE, USA; CVS Health/Aetna, Blue Bell, PA, USA; IQVIA, Falls Church, VA, USA

## Abstract

**Importance:** Active monitoring of health outcomes after COVID-19 vaccination provides early detection of rare outcomes that may not be identified in prelicensure trials.

**Objective:** To conduct near real-time monitoring of health outcomes following COVID-19 vaccination in the United States (US) pediatric population aged 6 months to 17 years.

**Design:** We evaluated 21 pre-specified health outcomes; 15 were sequentially tested through near real-time surveillance, and 6 were monitored descriptively within a cohort of vaccinated children. We tested for increased rate of each outcome following vaccination compared to a historical comparator cohort.

**Setting:** This population-based study was conducted under the US Food and Drug Administration public health surveillance mandate using three commercial claims databases.

**Participants:** Children aged 6 months to 17 years were included if they received a monovalent COVID-19 vaccine dose before early 2023 and had continuous enrollment in a medical health insurance plan from the start of an outcome-specific clean window to the COVID-19 vaccination dose.

**Exposure:** Exposure was defined as receipt of a monovalent BNT162b2, mRNA-1273, or NVX-CoV2373 COVID-19 vaccine dose. The primary analysis evaluated dose 1 and dose 2 combined, and secondary analyses evaluated each dose separately. Follow-up time was censored at death, disenrollment, end of risk window, end of study period, or a subsequent dose administration.

**Main Outcomes:** Twenty-one prespecified health outcomes.

**Results:** The study included 4,102,016 enrollees aged 6 months to17 years. Thirteen of 15 outcomes sequentially tested did not meet the threshold for a statistical signal. In the primary analysis, myocarditis or pericarditis signals were detected following BNT162b2 vaccine in children aged 12-17 years old and seizures/convulsions signals were detected following vaccination with BNT162b2 and mRNA-1273 in children aged 2-4/5 years. However, in a post-hoc sensitivity analysis, the seizures/convulsions signal was sensitive to background rates selection and was not observed when 2022 background rates were selected instead of 2020 rates.

**Conclusions and Relevance:** Of the two signaled outcomes, the myocarditis or pericarditis signals are consistent with previously published reports. The new signal detected for seizures/convulsions among younger children should be further investigated in a robust epidemiological study with better confounding adjustment.

## Key Points

### Question

Did active monitoring detect statistical signals for health outcomes following monovalent COVID-19 vaccination in the US children aged 6 months to 17 years?

### Findings

In this study including 4,102,106 vaccinated enrollees from three commercial claims databases, myocarditis or pericarditis signaled after BNT162b2 (12-17 years) and a new signal was detected for seizures/convulsions after BNT162b2 (2-4 years) and mRNA1273 COVID-19 vaccinations (2-5 years).

### Meaning

Near real-time monitoring of vaccines can rapidly identify potential safety concerns. While the myocarditis or pericarditis signal is consistent with existing evidence, the new seizures/convulsions signal should be interpreted cautiously given study limitations.

## Introduction

Three vaccines against COVID-19 are currently available for use in children in the United States (US) including the Pfizer-BioNTech (BNT162b2) and Moderna (mRNA-1273) COVID-19 mRNA vaccines for those aged 6 months to 17 years and the protein-based Novavax COVID-19 vaccine (NVX-CoV2373) for those aged 12 to 17 years.^1–3^ As of May 2023, the Centers for Disease Control and Prevention (CDC) reported 31.78 million children had received at least one COVID-19 vaccine dose and 26.2 million children had completed the primary series out of approximately 73 million children aged 6 months to 17 years in the US.^4^ The US Food and Drug Administration (FDA), utilizing the Biologics Effectiveness and Safety (BEST) Initiative, has been monitoring the safety of COVID-19 vaccines in children since their authorization by applying a near real-time monitoring framework to evaluate the safety of COVID-19 vaccines. This process is a signal detection or screening method and the first step in safety monitoring of these vaccines. This framework is designed to be sensitive enough to rapidly detect less common safety signals. However, results of such study design do not establish a causal relationship between the vaccines and health outcomes and need to be interpreted with caution because of the limited adjustment for confounding and other forms of bias.

The BEST analysis of COVID-19 vaccines in children initially focused on the BNT162b2 COVID-19 monovalent vaccine authorized for use in children aged 5 to 17 years. Surveillance has since been expanded as additional pediatric age groups and vaccine brands received authorization through late 2022. Results of the initial, more limited, safety surveillance in children have been previously published.^5^ In this report, we present results from the expanded monitoring of health outcomes in children after exposure to the ancestral monovalent COVID-19 vaccines in the US targeting the original COVID-19 strain.

## Methods

### Study Objective

This study evaluated 21 pre-specified health outcomes after exposure to BNT162b2, mRNA-1273, or NVX-CoV2373 monovalent COVID-19 vaccines in children 6 months to 17 years old by applying a near real-time monitoring framework using health care data from three commercial claims databases in the US. Fifteen outcomes underwent sequential testing, and 6 outcomes were only monitored descriptively due to lack of historical rates.

### Data Sources

The study used commercial administrative claims data from Optum, Carelon Research, and CVS Health containing longitudinal medical and pharmacy claims data supplemented with vaccination data from participating local and state Immunization Information Systems (IIS) (Supplemental Table 1).^6^

### Study Population and Period

The study included pediatric enrollees aged 6 months to 17 years who received a monovalent COVID-19 vaccine from the earliest date of its Emergency Use Authorization by age group through April 2023 (Optum), March 2023 (Carelon Research), and February 2023 (CVS Health) (Supplemental Table 2). The surveillance concluded on these respective dates because of limited accrual of exposures and health outcomes. The sequential analyses for most outcomes did not reach the expected number of events that was pre-specified based on anticipated vaccination uptake in a 6-month period post-authorization of individual vaccine products. Another contributing factor to halting surveillance was the de-authorization of the original monovalent mRNA-1273 and BNT162b2 doses for persons of all ages on April 18, 2023.^7^

The inclusion criteria for the study included enrollment on the vaccination date and continuous enrollment in a participating medical health insurance plan from the start of an outcome-specific clean window to the COVID-19 vaccination date so that only new incident diagnosis of an outcome in the post-vaccination risk window would contribute to the analysis (Supplemental Table 3).

### Exposures and Follow Up

Exposure was defined as the receipt of a BNT162b2, mRNA-1273, or NVX-CoV2373 monovalent COVID-19 vaccine dose identified using brand and dose-specific Current Procedural Terminology/ Healthcare Common Procedure Coding System codes^8^ and National Drug Codes (Supplemental Table 4). Dose number was assigned based on the chronological order in which vaccinations were observed since administration codes were not available in pharmacy claims. The primary analyses included all follow-up time accrued after dose 1 and dose 2 combined. Secondary analyses included stratification by dose number and follow-up time accrued after the individual dose (including monovalent third/booster doses) through censoring at subsequent vaccination, death, disenrollment, end of risk window, or end of study period.

### Health Outcomes

Twenty-one pre-specified health outcomes were defined using claims-based algorithms^9–10^. Outcomes were selected through clinical consultation and literature review. Fifteen health outcomes were assessed using sequential testing comparing rates to historical outcome rates (additional information on historical rates below), and 6 were only monitored descriptively due to lack of historical rates.^10^ The outcome myocarditis, pericarditis, or co-occurring myocarditis and pericarditis (hereafter referred to as myocarditis/pericarditis) was assessed using four different definitions with varying risk windows and care settings (included in Supplemental Table 3) based on evidence from prior surveillance efforts and clinical input.

### Descriptive Monitoring

We estimated outcome rates in the vaccinated population stratified by age, sex, region, urban/rural status, data source, and vaccine brand on a monthly basis.

### Sequential Testing

Monthly sequential testing was conducted using the Poisson Maximized Sequential Probability Ratio Test (PMaxSPRT) to detect statistical signals by generating the incidence rate ratio (IRR) comparing outcome rates following vaccination to database-specific historical (expected) rates for 15 health outcomes.^11^

We estimated annual historical rates for 2019 and 2020 as well as during the COVID-19 pandemic between April and December 2020. Historical rates were adjusted for claims processing delay to account for observation delay and standardized by age and sex where case counts permitted.^12^ Selection of the historical comparator rate was based on the overlap between the 95% confidence intervals (CI) for the periods prior to and during the COVID-19 pandemic. If rates in these two historical periods differed substantially, we selected either the minimum or more stable rate as a more conservative approach.

Tests were stratified by age based on age-group-specific authorizations by vaccine brand as well as availability of background rates using historical comparator data. For BNT162b2, this included ages 6 months-4 years, 6 months-1 year and 2-4 years (for seizure/convulsions only), 5-11 years, 12-15 years, and 16-17 years. For mRNA-1273, this included 6 months-5 years, 6 months-1 year and 2-5 years (for seizure or convulsions only), 6-11 years, 12-15 years, and 16-17 years. For NVX-CoV2373, the age groups were 12-15 years and 16-17 years.

Sequential testing for each outcome commenced when a minimum of three cases accrued in the risk window. One-tailed tests were used with the null hypothesis that the observed rate was no greater than the historical comparator rate beyond a pre-specified test margin for each outcome-dose-age group being sequentially tested, with an alpha level of 1%. A stringent alpha level was selected to increase the specificity of signals detected from testing multiple outcomes across different analyses. The log likelihood ratio was calculated comparing the likelihood of the observed IRR and the null hypothesis.

At each test, if the log likelihood ratio exceeded a pre-specified critical value, the null hypothesis was rejected, and a signal was declared. Surveillance continued until a signal was detected, the pre-specified maximum surveillance length was reached, or the end of study period was reached.^10^

### Signal Characterization

Signal characterization was conducted after a signal was identified to provide data quality assessment.^13^ We conducted quality checks to rule out database errors or changes in the patterns of diagnosis codes used to identify events in the study period; estimated the relative risk of outcomes within demographic strata by age and sex; examined the timing of outcomes occurrence during the pre-specified risk window; and assessed whether the signal was sensitive to changes in background rates selection by conducting a post-hoc sensitivity analysis using the updated 2022 background rates as the historical comparator in sequential testing.

### Medical Record Review

Medical record review was conducted for the myocarditis/pericarditis outcome following identification of a signal. Brighton Collaboration case definitions were used to adjudicate cases.^14^ Records meeting the confirmed or probable Brighton classifications were considered true myocarditis/pericarditis cases for the validation analyses.

## Results

### Descriptive Monitoring

A total of 8,444,355 monovalent COVID-19 vaccine doses were administered to 4,102,016 enrollees aged 6 months -17 years (Figure 1). This included 8,121,591 BNT162b2 (Dose 1: 3,843,778, Dose 2: 3,235,442, Dose 3/ Monovalent Booster: 1,033,036 and Unknown/Unclear: 9,335), and 322,628 mRNA-1273 doses (Dose 1: 173,857, Dose 2: 140,734, Dose 3/Monovalent Booster: 5,284 and Unknown/Unclear: 2,753) administered to children aged 6 months-17 years, as well as 136 NVX-CoV2373 doses (Dose 1: 63, Dose 2: 43, Dose 3/Monovalent Booster and Unknown/Unclear: 30) administered to children aged 12-17 years (Table 1a). Demographic characteristics of the vaccinated population except age reported at first dose were largely similar across vaccine brands. While majority (93.4%) of BNT162b2-vaccinated individuals were 5 years old and older; majority (97.6%) of mRNA-1273-vaccinated individuals were younger than 5 years old. Across all vaccine brands, 2,058,142 (50.2%) were males and 3,901,370 (95.2%) lived in an urban area (Table 1b). We observed low case counts for outcomes that were monitored descriptively only (<80 cases of any given outcome in all three databases combined) (Supplemental Table 5).

**Figure 1.**
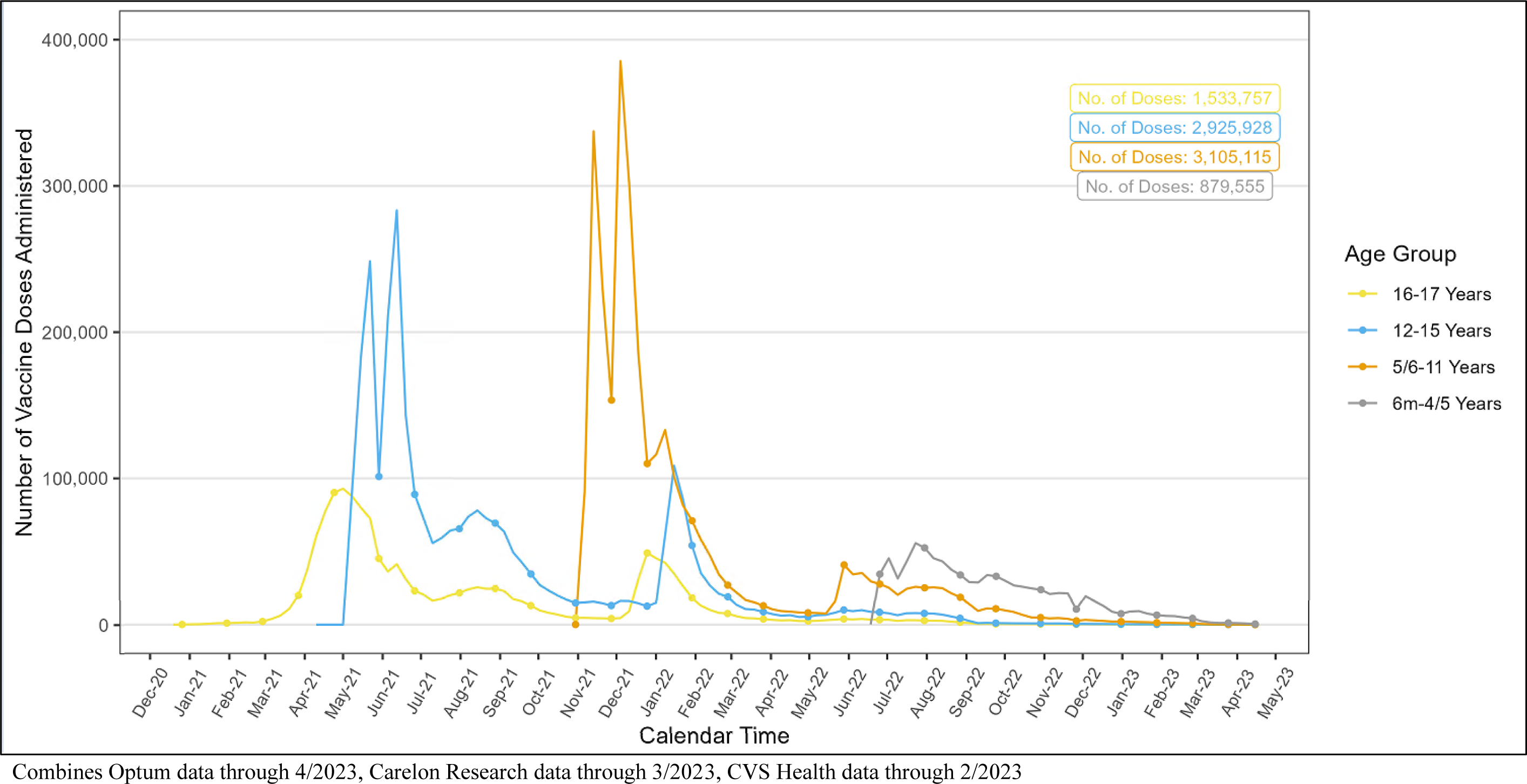
Distribution of Monovalent Doses of BNT162b2, mRNA-1273, and NVX-CoV2373 COVID-19 Vaccines Administered Over Time by Age Group, All Data Sources.

**Table 1a.** Vaccine Dose Counts for Monovalent BNT162b2, mRNA-1273, NVX-CoV2373 COVID-19 Vaccines Administered to the Pediatric Population, All Data Sources^a^.

| Vaccine Dose | BNT162b2 |  | mRNA-1273 |  | NVX-CoV2373 |  |
| --- | --- | --- | --- | --- | --- | --- |
|  | N | % | N | % | N | % |
| **Total** | 8,121,591 | 100 | 322,628 | 100 | 136 | 100 |
| **Dose 1** | 3,843,778 | 47.33 | 173,857 | 53.89 | 63 | 46.32 |
| **Dose 2** | 3,235,442 | 39.84 | 140,734 | 43.62 | 43 | 31.62 |
| **Dose 3/Monovalent Booster** | 1,033,036 | 12.72 | 5,284 | 1.64 | § | § |
| **Unknown/Unclear** | 9,335 | 0.11 | 2,753 | 0.85 | § | § |

<sup>a</sup> Combines Optum data through 4/2023, Carelon Research data through 3/2023, CVS Health data through 2/2023
§ Cell sizes 1-10 and cells that can be used to back-calculate small cell sizes were masked for confidentiality

**Table 1b.**
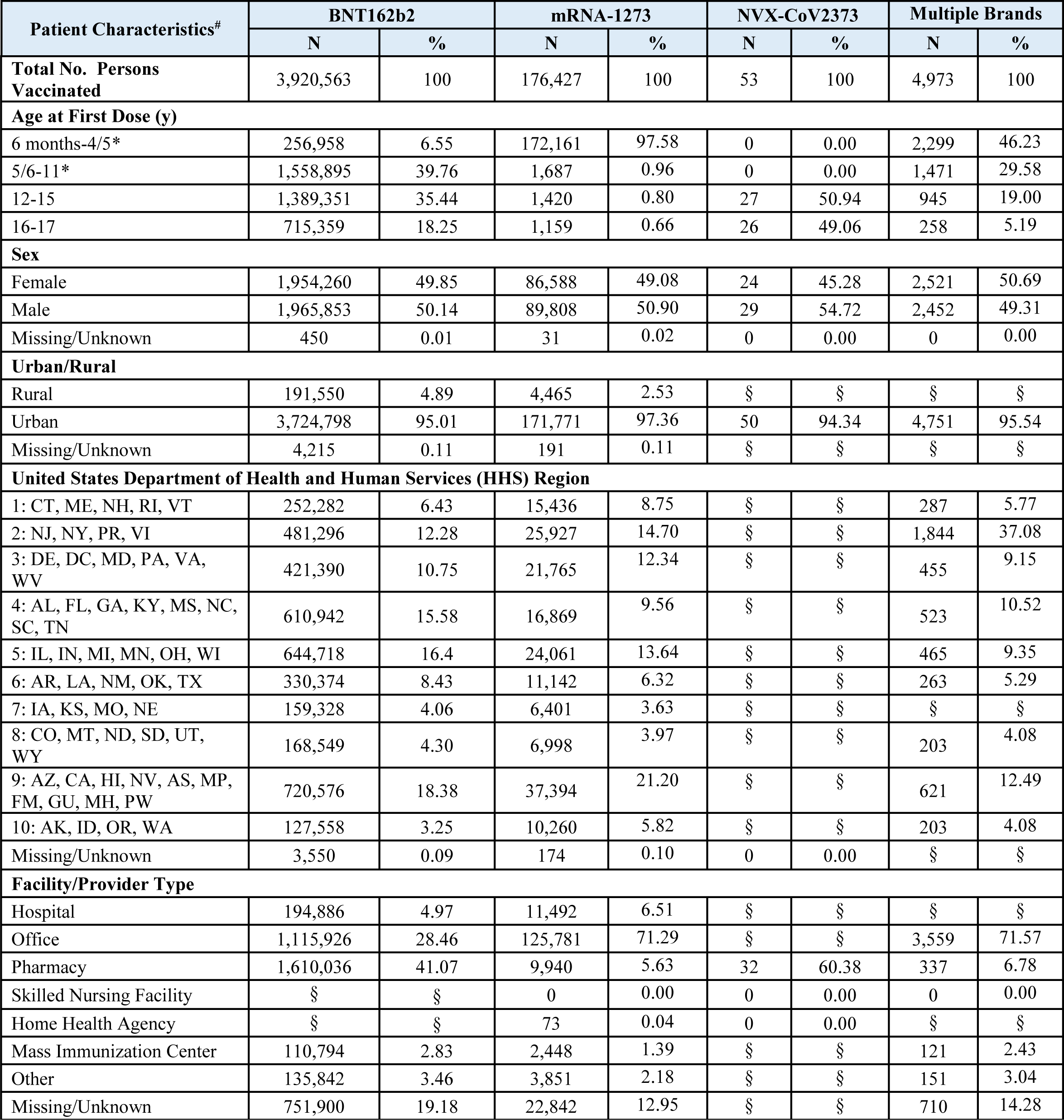
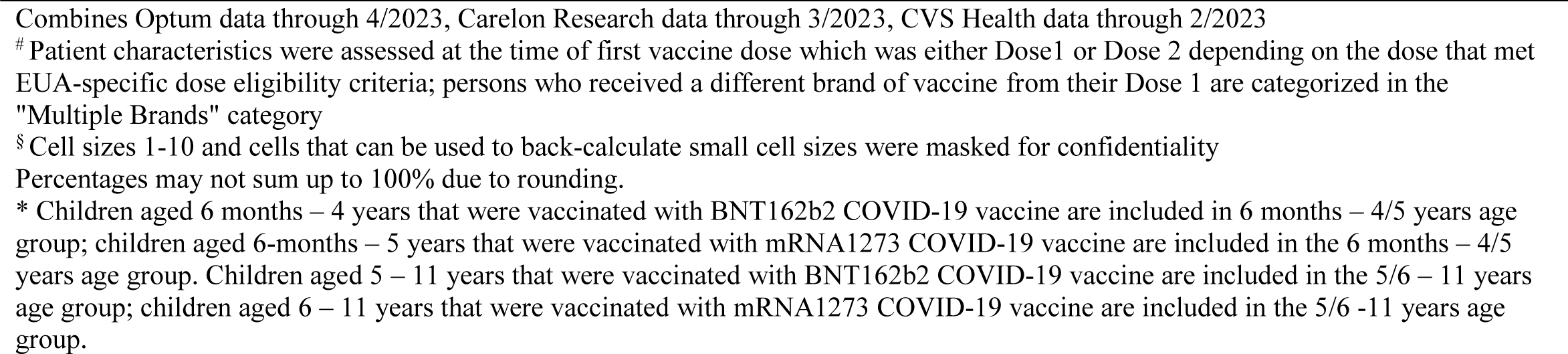
Characteristics of Pediatric Population Receiving Monovalent BNT162b2, mRNA-1273, and NVX-CoV2373 COVID-19 Vaccines, All Data Sources.

| Patient Characteristics<sup>#</sup> | BNT162b2 |  | mRNA-1273 |  | NVX-CoV2373 |  | Multiple Brands |  |
| --- | --- | --- | --- | --- | --- | --- | --- | --- |
|  | N | % | N | % | N | % | N | % |
| **Total No. Persons Vaccinated** | 3,920,563 | 100 | 176,427 | 100 | 53 | 100 | 4,973 | 100 |
| **Age at First Dose (y)** |  |  |  |  |  |  |  |  |
| 6 months-4/5* | 256,958 | 6.55 | 172,161 | 97.58 | 0 | 0.00 | 2,299 | 46.23 |
| 5/6-11* | 1,558,895 | 39.76 | 1,687 | 0.96 | 0 | 0.00 | 1,471 | 29.58 |
| 12-15 | 1,389,351 | 35.44 | 1,420 | 0.80 | 27 | 50.94 | 945 | 19.00 |
| 16-17 | 715,359 | 18.25 | 1,159 | 0.66 | 26 | 49.06 | 258 | 5.19 |
| **Sex** |  |  |  |  |  |  |  |  |
| Female | 1,954,260 | 49.85 | 86,588 | 49.08 | 24 | 45.28 | 2,521 | 50.69 |
| Male | 1,965,853 | 50.14 | 89,808 | 50.90 | 29 | 54.72 | 2,452 | 49.31 |
| Missing/Unknown | 450 | 0.01 | 31 | 0.02 | 0 | 0.00 | 0 | 0.00 |
| **Urban/Rural** |  |  |  |  |  |  |  |  |
| Rural | 191,550 | 4.89 | 4,465 | 2.53 | § | § | § | § |
| Urban | 3,724,798 | 95.01 | 171,771 | 97.36 | 50 | 94.34 | 4,751 | 95.54 |
| Missing/Unknown | 4,215 | 0.11 | 191 | 0.11 | § | § | § | § |
| **United States Department of Health and Human Services (HHS) Region** |  |  |  |  |  |  |  |  |
| 1: CT, ME, NH, RI, VT | 252,282 | 6.43 | 15,436 | 8.75 | § | § | 287 | 5.77 |
| 2: NJ, NY, PR, VI | 481,296 | 12.28 | 25,927 | 14.70 | § | § | 1,844 | 37.08 |
| 3: DE, DC, MD, PA, VA, WV | 421,390 | 10.75 | 21,765 | 12.34 | § | § | 455 | 9.15 |
| 4: AL, FL, GA, KY, MS, NC, SC, TN | 610,942 | 15.58 | 16,869 | 9.56 | § | § | 523 | 10.52 |
| 5: IL, IN, MI, MN, OH, WI | 644,718 | 16.4 | 24,061 | 13.64 | § | § | 465 | 9.35 |
| 6: AR, LA, NM, OK, TX | 330,374 | 8.43 | 11,142 | 6.32 | § | § | 263 | 5.29 |
| 7: IA, KS, MO, NE | 159,328 | 4.06 | 6,401 | 3.63 | § | § | § | § |
| 8: CO, MT, ND, SD, UT, WY | 168,549 | 4.30 | 6,998 | 3.97 | § | § | 203 | 4.08 |
| 9: AZ, CA, HI, NV, AS, MP, FM, GU, MH, PW | 720,576 | 18.38 | 37,394 | 21.20 | § | § | 621 | 12.49 |
| 10: AK, ID, OR, WA | 127,558 | 3.25 | 10,260 | 5.82 | § | § | 203 | 4.08 |
| Missing/Unknown | 3,550 | 0.09 | 174 | 0.10 | 0 | 0.00 | § | § |
| **Facility/Provider Type** |  |  |  |  |  |  |  |  |
| Hospital | 194,886 | 4.97 | 11,492 | 6.51 | § | § | § | § |
| Office | 1,115,926 | 28.46 | 125,781 | 71.29 | § | § | 3,559 | 71.57 |
| Pharmacy | 1,610,036 | 41.07 | 9,940 | 5.63 | 32 | 60.38 | 337 | 6.78 |
| Skilled Nursing Facility | § | § | 0 | 0.00 | 0 | 0.00 | 0 | 0.00 |
| Home Health Agency | § | § | 73 | 0.04 | 0 | 0.00 | § | § |
| Mass Immunization Center | 110,794 | 2.83 | 2,448 | 1.39 | § | § | 121 | 2.43 |
| Other | 135,842 | 3.46 | 3,851 | 2.18 | § | § | 151 | 3.04 |
| Missing/Unknown | 751,900 | 19.18 | 22,842 | 12.95 | § | § | 710 | 14.28 |

Combines Optum data through 4/2023, Caredon Research data through 3/2023, CVS Health data through 2/2023

### Sequential Testing

Among 15 outcomes that were sequentially tested, two outcomes met the statistical threshold for a signal including myocarditis/pericarditis in ages 12-15 and 16-17 years, and seizures/convulsions in ages 2-4/5 years.

Myocarditis/pericarditis signaled for all definitions in the primary analysis following BNT162b2 COVID-19 vaccination among children aged 12-15 and 16-17 years in all three databases. Additionally, dose-specific signals for one or more definitions of the outcomes were detected in ages 12-17 years following dose 1, dose 2, and dose 3 of BNT162b2 vaccine in at least one of the three databases (Table 2).

**Table 2.**
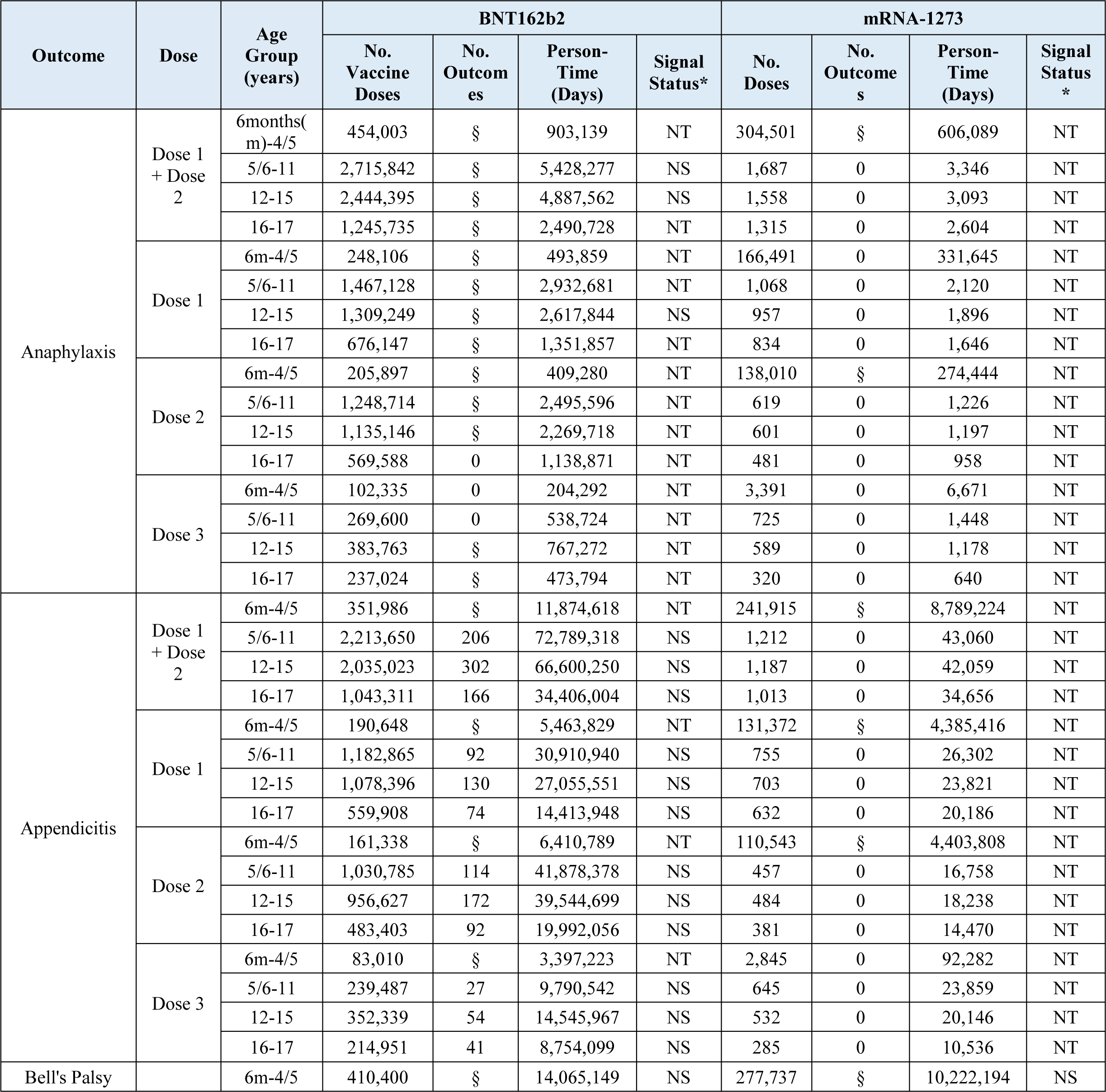
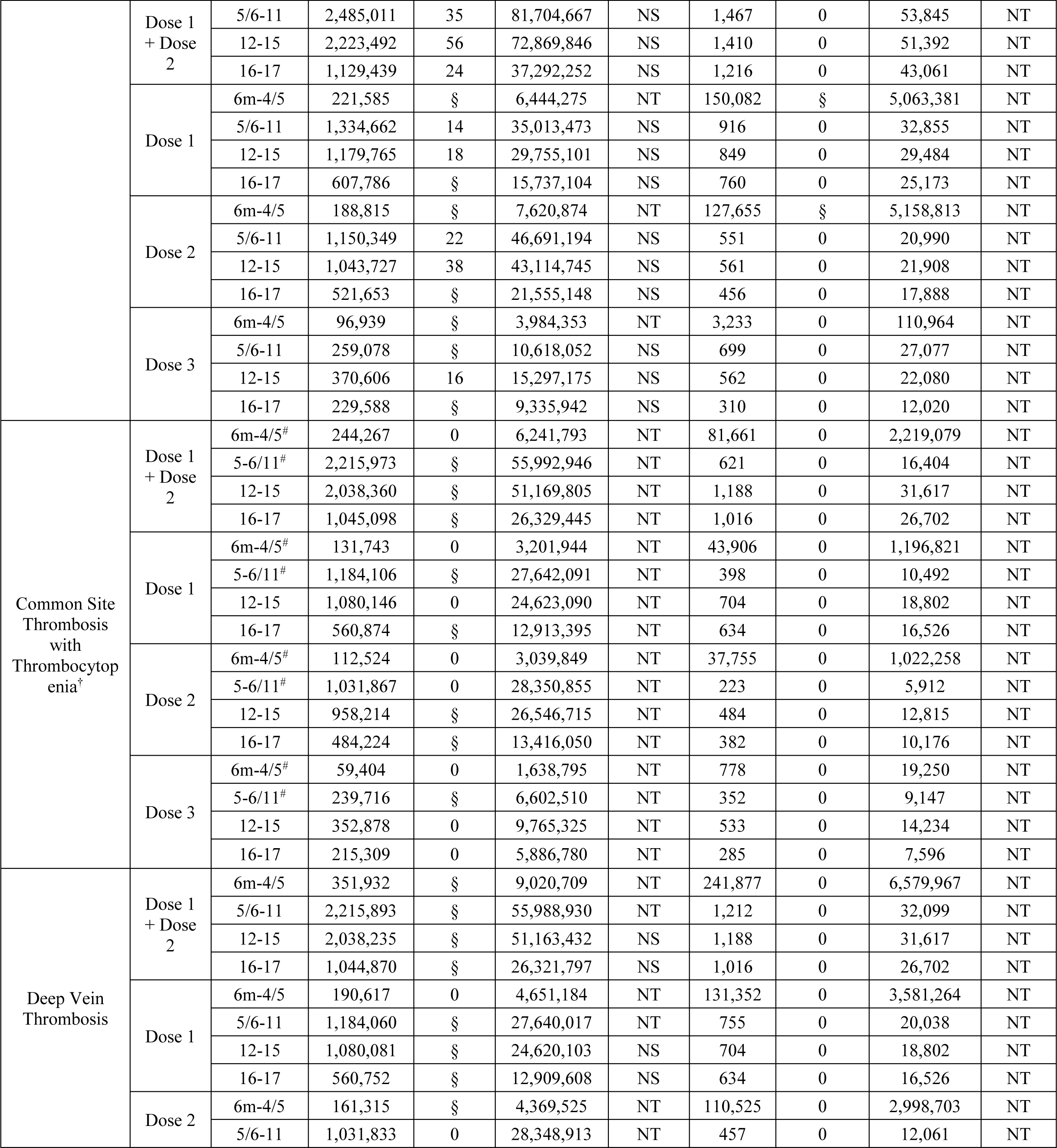
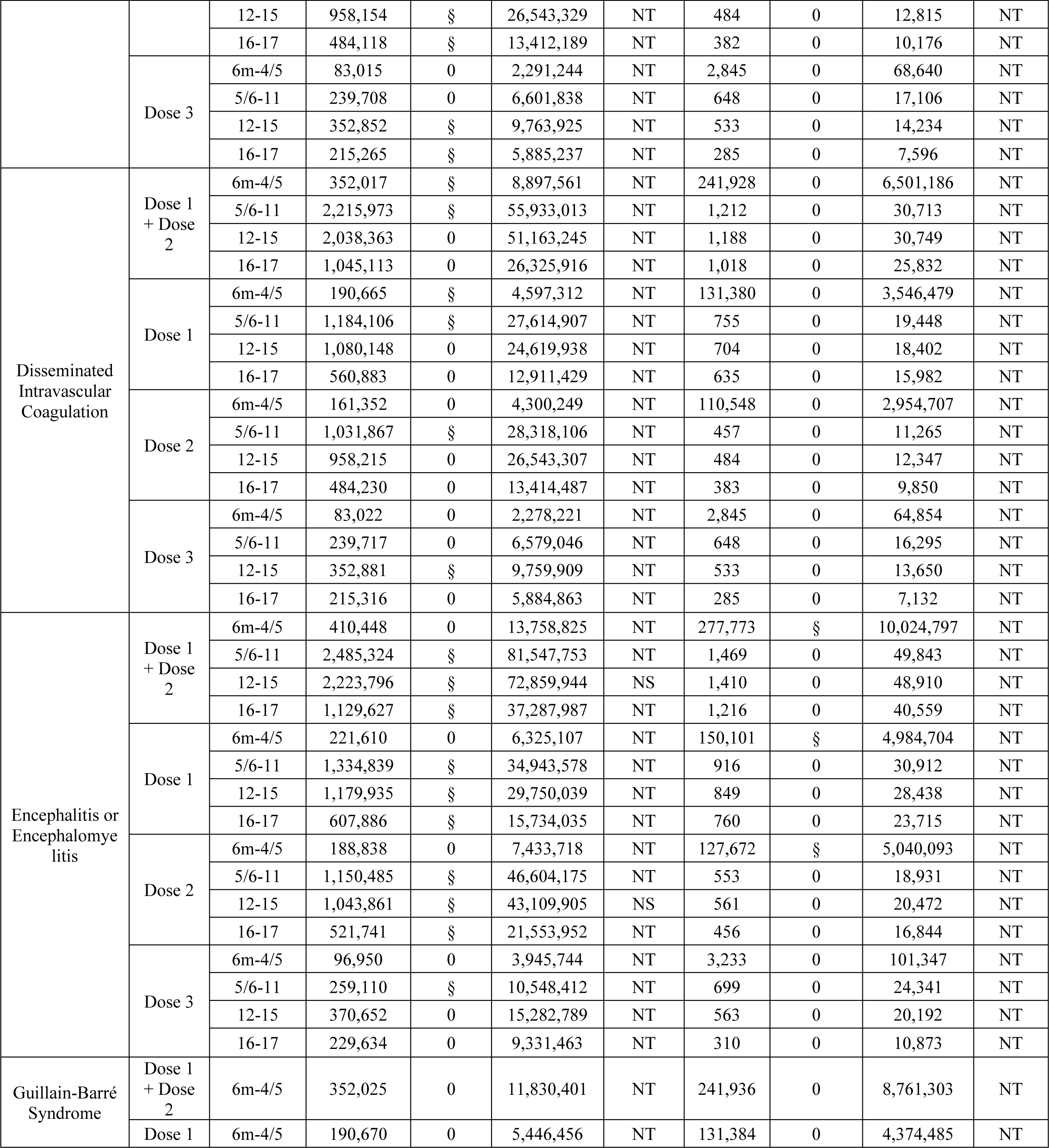
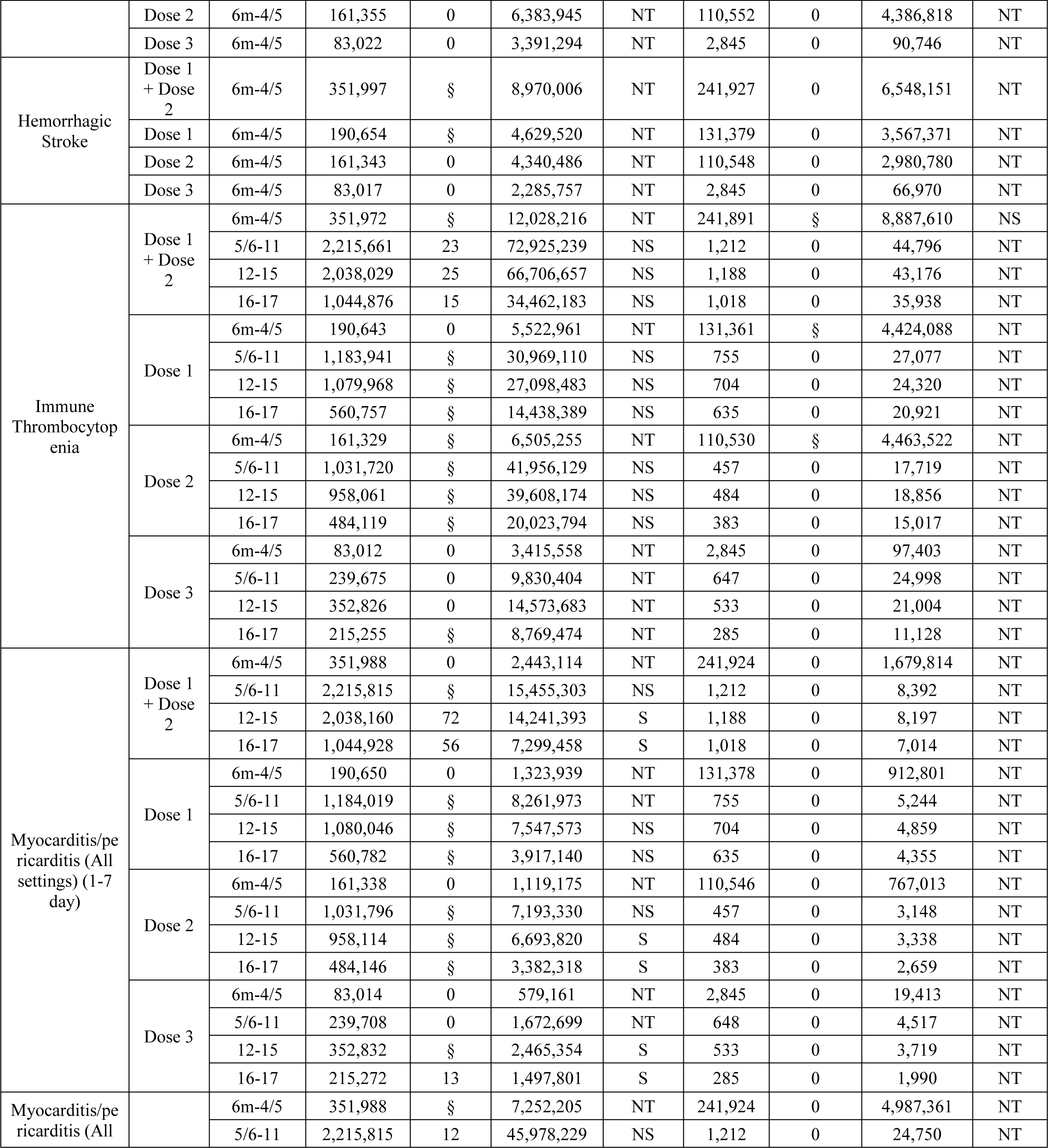
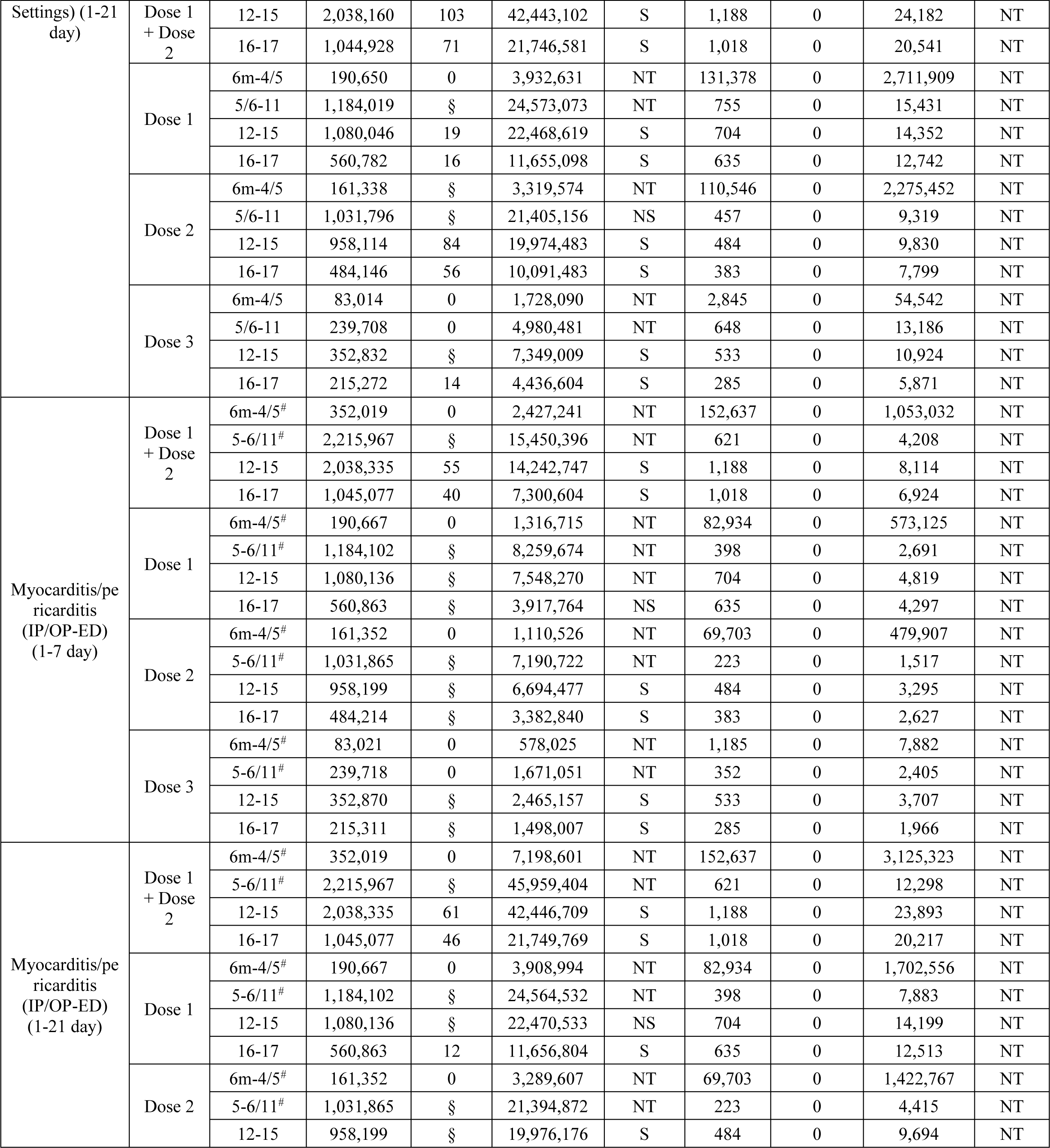
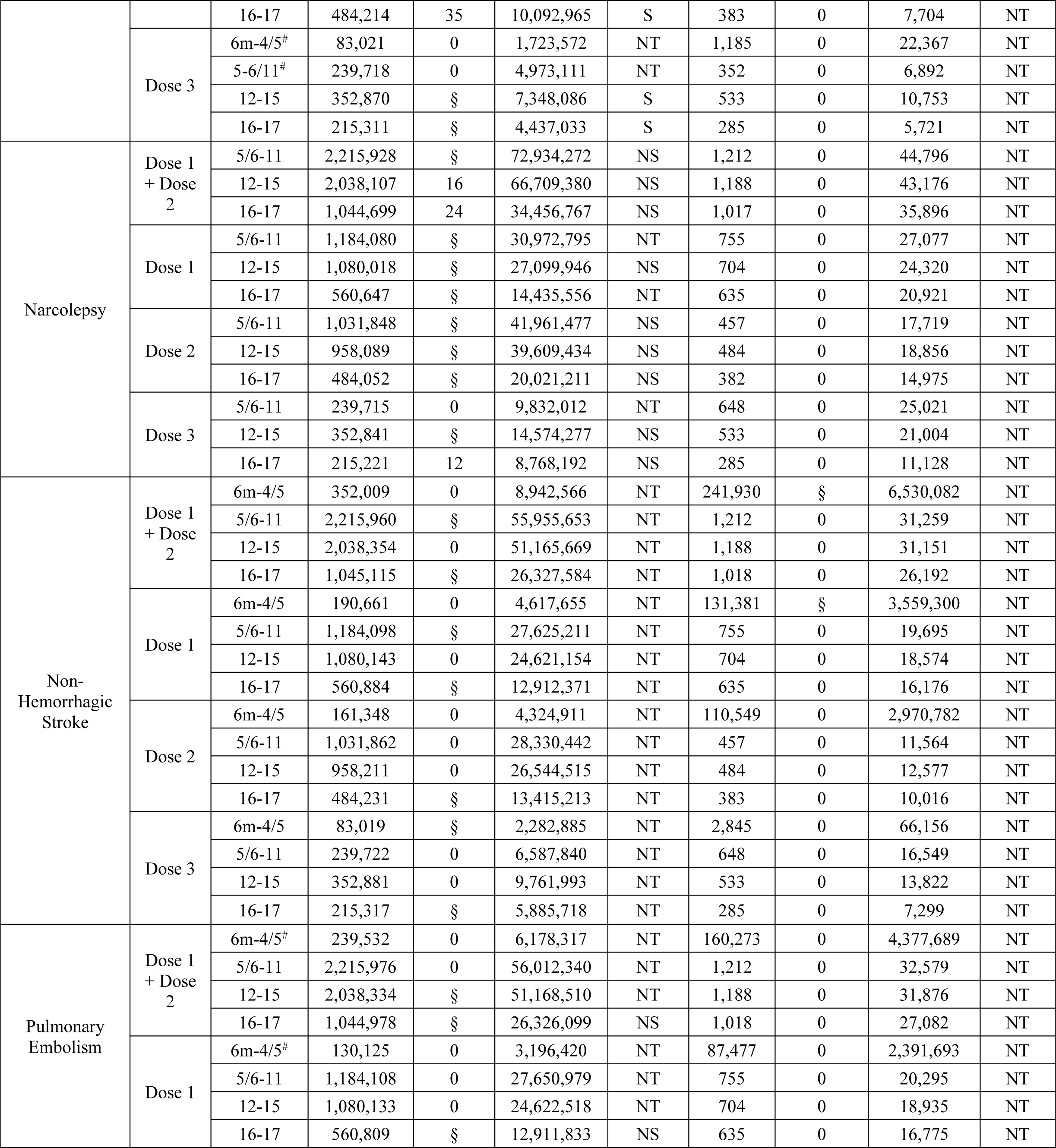
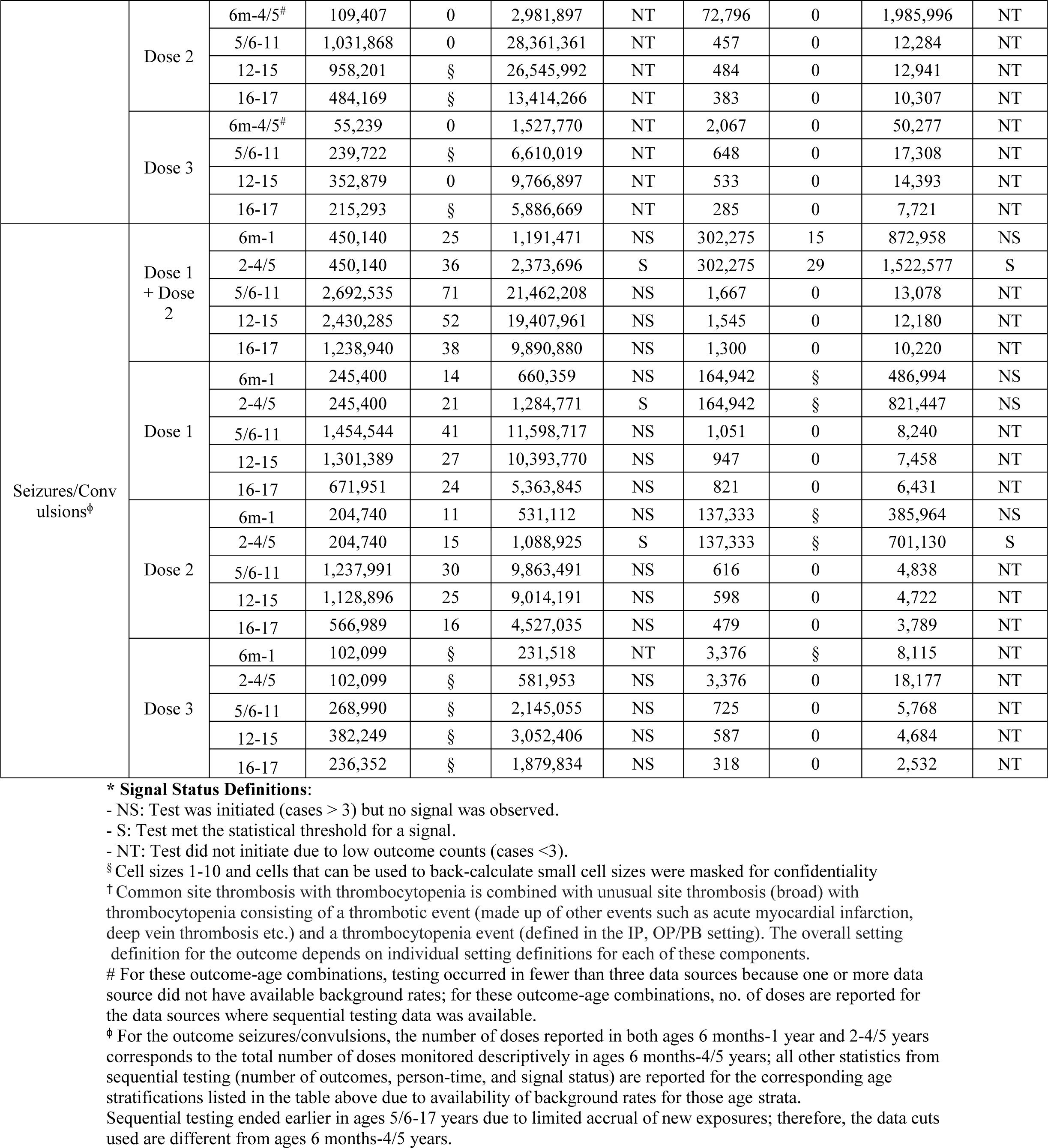
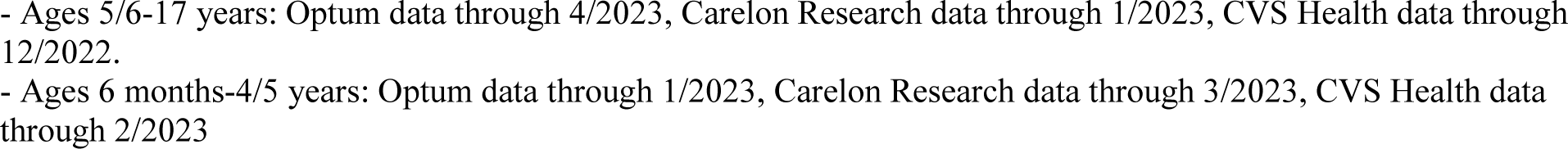
Sequential Testing Results for the Pediatric Population Receiving BNT162b2 or mRNA-1273 Monovalent COVID-19 Vaccines Stratified by Vaccine dose and Outcome.

In the primary analysis, seizures/convulsions met the statistical threshold for a signal in children aged 2-4 years following BNT162b2 vaccination in all three databases, and in children aged 2-5 years following mRNA-1273 vaccination in two of the three databases. Dose-specific signals for seizures/convulsions were detected in two of the three databases following dose 1 and dose 2 BNT162b2 vaccination in ages 2-4 years and following dose 2 mRNA-1273 vaccination in ages 2-5 years (Table 2).

Sequential testing did not initiate for any of the 15 outcomes for mRNA-1273 in ages 5-17 years and NVX-CoV2373 in ages 12-17 years, because no outcomes were observed in any databases.

### Signal Characterization

Since myocarditis/pericarditis is a known adverse outcome following COVID-19 mRNA vaccinations, further signal characterization activities were not conducted. In the evaluation of the seizures/convulsions signal among children aged 2-4/5 years, none of the pre-specified data quality checks, including claims duplication, timing of events within pre-specified risk window, and IRR estimates, raised any data quality concerns.

There were 72 observed seizures/convulsions cases among children aged 2-4/5 years and >50% of these cases met the definition of febrile seizures. No differences in rates of seizures/convulsions by sex were identified. The timing of cases did not indicate substantial clustering with cases distributed across the 0-7-day risk window; 31.9% of the seizures/convulsions cases occurred within the 0-1 days following COVID-19 vaccination. The median time between vaccination and diagnosis of seizures/convulsions was 2 days. (Supplemental Figure 1).

The seizures/convulsions signal was sensitive to changes in the selection of comparator rates. Evaluation of the annual background rate of seizures/convulsions indicated that the rates used in the primary analyses (2020) were substantially lower than rates in 2022. Background rates in 2022 ranged from approximately 2.2-2.4 times the 2020 rates across three databases (Supplemental Table 6). A post-hoc sensitivity analysis using 2022 background rates as the comparator in sequential testing did not identify any seizures/convulsions signals.

### Medical Record Review

Of the 153 cases of myocarditis/pericarditis after COVID-19 vaccination among children aged 12-17 years, medical record review was conducted for a sample of 40 cases whose records could be obtained. Twenty-nine of these cases (72.5%) were confirmed as true cases of myocarditis/pericarditis, of which 27 patients were male, and 19 were hospitalized with a median length of hospital stay of 2 days (interquartile range: 1, 3). The median time from vaccination to presentation of myocarditis/pericarditis event was 3 days (interquartile range: 2, 5). Medical record review and further validation efforts for seizures/convulsions are currently underway.

## Discussion

Our near real-time monitoring of 21 pre-specified health outcomes following monovalent COVID-19 vaccines detected signals for myocarditis/pericarditis in the age group 12-17 years and seizures/convulsions in the age group 2-4/5 years. We did not detect signals for other outcomes that were sequentially tested.

The myocarditis/pericarditis signal is consistent with peer-reviewed publication reports demonstrating an elevated risk of this outcome following mRNA vaccines among younger males aged 12 to 29 years.^15–17^ Myocarditis/pericarditis is a rare event with a reported average incidence of 39.3 cases per one million vaccine doses administered in children aged 5-17 years within 7 days after BNT162b2 vaccination.^18–19^ We did not detect a signal for myocarditis/pericarditis in children younger than 12 years old which is consistent with reports from other surveillance systems.^20–21^

The seizures/convulsions signal in children aged 2-4/5 years has not been previously reported for this age group in active surveillance studies of mRNA COVID-19 vaccines. However, there are reports from the Vaccine Adverse Events Reporting System (VAERS) database which is a passive reporting system and has limitations. In an analysis of VAERS data, only 8 seizures were identified following approximately one million mRNA vaccinations through August 2022 in the age group 6 months to 5 years. Six of the 8 seizures were afebrile on medical evaluation.^22^ Pre-licensure vaccine safety data indicate that among young children, seizures/convulsions following mRNA COVID-19 vaccines are rare; clinical trial of 3,013 BNT162b2 vaccine recipients in children aged 6 months to 4 years reported only 5 febrile convulsions cases and only one of those (in a 6-month-old participant) was considered possibly related to the vaccine or may also have been caused by a concurrent viral infection.^23^ Among studies conducted in children with childhood epilepsy (<18 years), no increased risk of medically attended seizures was identified following immunization with COVID-19 vaccines. Seizure risk after COVID-19 vaccination was lower in children who were seizure free for more than six months before vaccination. However, the incidence of general adverse events after vaccination was low with no severe adverse events recorded. ^24–25^ Generally, there is limited evidence linking the mRNA COVID-19 vaccines to a seizure onset among vaccinated children aged 2-4/5 years.

The new seizures/convulsions signal observed in our study should be interpreted with caution and further investigated in a more robust epidemiological study. Our study utilized a broad seizures/convulsions outcome definition with a 0-7-day risk window because of its applicability to older children. However, in children younger than 5 years, vaccine-related seizures typically manifest as a febrile seizure.^26^ Although the majority of seizures/convulsions cases met the febrile seizure definition, there was no statistically significant clustering observed at days 0-1. Since febrile seizures can be common in young children for a variety of reasons; the analysis may have identified febrile seizures unrelated to the vaccination later in the risk window.

The post-hoc sensitivity analyses using the 2022 background rates as comparators in sequential testing did not yield any seizures/convulsions signals which suggest that our results are sensitive to comparator rate selection. The decision to use 2020 seizures/convulsions rates as comparators was to maximize sensitivity in the primary analysis. However, seizures/convulsions rates in this age group in 2022 were twice as high as 2020 rates. There could be a couple of potential reasons for elevated outcome rates in 2022 compared to 2020. First, there was an increased incidence of respiratory infections (influenza and respiratory syncytial virus) which are shown to be associated with febrile seizure in younger children, during the study period (mid-2022 to mid-2023) compared to 2020.^27–30^ Second, in 2020, there were likely fewer emergency department visits for seizure-related events compared to 2022 because of COVID-19 pandemic healthcare resource limitations.^31^

Our study has a number of strengths. First, the study included a large, geographically diverse population from three US commercial health insurance databases. Due to availability of more complete information from claims supplemented with IIS data and a short data lag from health encounters, we monitored monovalent COVID-19 vaccines safety in a near real-time manner. Additionally, a subset of the identified myocarditis/pericarditis cases were confirmed through medical record review.

The study also has some limitations. We used a near–real-time surveillance method which may be sensitive to comparator rate selection and does not include controlling for bias and confounding. Therefore, results from this study do not establish a causal relationship between the vaccines and health outcomes, and signals should be further evaluated. Secondly, this study only includes data from a commercially insured pediatric population and may not be nationally representative. Furthermore, the study may have limited power to detect small increases in risk of outcomes in certain subgroups or in the cases of more recently authorized vaccines, such as NVX-CoV2373 in children aged 12-17 years as well as mRNA-1273 in those aged 5-17 years.

## Conclusion

Our study detected a signal for myocarditis/pericarditis in older children which is consistent with existing literature, and a new signal for seizures/convulsions in young children that is being further evaluated in a more robust study. FDA concludes that the known and potential benefits of COVID-19 vaccination outweigh the known and potential risks of COVID-19 infection. This study was conducted under the FDA BEST Initiative which plays a major role in the larger US federal government vaccine safety monitoring efforts and further supports regulatory decision-making regarding COVID-19 vaccines.

## Data Availability

The study protocol was publicly posted previously and a brief report was published describing the preliminary results of BNT162b2 COVID-19 Vaccine in Children Aged 5 to 17 Years on JAMA Pediatrics, as referenced in the manuscript. Data analyses and related documents can be made available where needed, by contacting the corresponding author. De-identified participant data will not be shared without approval from the data partners.

https://covid.cdc.gov/covid-data-tracker

## Additional Contributions

We thank Tainya C. Clarke, Sylvia Cho and Carla E. Zelaya of the US Food and Drug Administration; Anchi Lo, Bing Lyu, Nimesh Shah, Derek Phan, Gyanada Acharya, Kamakshi Sirpal, Laurie Feinberg, Lloyd Marks, Minisha Kochar, Sandia Akhtar, Shruti Parulekar, Stella Zhu, William (Trey) Minter, Yeerae Kim, Yixin Jiao, and Zhiruo (Cassie) Wan of Acumen; Nancy B Shaik, Ana M Martinez-Baquero, Smita Bhatia, Vaibhav Sharma, Carla Brannan of CVS Health and Charlalynn Harris and Danielle Cooper formerly CVS Health; Shiva Vojjala, Ramya Avula, Shiva Chaudhary, Shanthi P Sagare, Ramin Riahi, Navyatha Namburu, and Grace Stockbower of Carelon Research; Michael Goodman, Michael Bruhn, and Ruth Weed of IQVIA; Grace Yang, Sarah Sargen, Alexandra Stone, Wafa Tarazi, Megan Ketchell, Kathryn Federici, Amaka Ume, Emily Myers, Eli Wolter, Jackson Slaney, Bobby Smith, Lauren Peetluk, and Elizabeth Bell of Optum.

## References

1. FDA. (2022, June 30). Covid-19 Vaccines. U.S. Food and Drug Administration. Retrieved from http://www.fda.gov/emergency-preparedness-and-response/coronavirus-emergency-preparedness-and-response/coronavirus-disease-2019-covid-19/covid-19-vaccines#authorized-vaccines

2. Centers for Disease Control and Prevention. (2022, June 18). CDC Recommends COVID-19 Vaccines for Young Children. Centers for Disease Control and Prevention. Retrieved from https://www.cdc.gov/media/releases/2022/s0618-children-vaccine.html

3. U.S. Food and Drug Administration. (2022, August 19). Covid-19 Vaccines. Retrieved August 24, 2022, from https://www.fda.gov/media/159902/download

4. Centers for Disease Control and Prevention. COVID Data Tracker. Atlanta, GA: UUUU.S. Department of Health and Human Services, CDC; 2023, July 11. https://covid.cdc.gov/covid-data-tracker

5. Hu M, Wong HL, Feng Y, et al. Safety of the BNT162b2 COVID-19 Vaccine in Children Aged 5 to 17 Years. JAMA Pediatr. Published online May 22, 2023. Doi:10.1001/jamapediatrics.2023.1440

6. Centers for Disease Control and Prevention. (2019, June 7). Immunization Information Systems (IIS). Centers for Disease Control and Prevention. Retrieved from https://www.cdc.gov/vaccines/programs/iis/index.html

7. Commissioner, O. Coronavirus (COVID-19) update: FDA authorizes changes to simplify use of bivalent mrna COVID-19 vaccines, U.S. Food and Drug Administration. Retrieved from https://www.fda.gov/news-events/press-announcements/coronavirus-covid-19-update-fda-authorizes-changes-simplify-use-bivalent-mrna-covid-19-vaccines

8. License for use of current procedural terminology, Fourth edition (‘Cpt®’) CMS.gov. Available at: https://www.cms.gov/license/ama?file=%2Ffiles%2Fzip%2Flist-codes-effective-january-1-2023-published-december-1-2022.zip (Accessed: 31 August 2023).

9. Center for Biologics Evaluation and Research, BEST Initiative. COVID-19 vaccine safety protocol supplemental. Published 2021. https://bestinitiative.org/vaccines-and-allergenics

10. Center for Biologics Evaluation and Research, BEST Initiative (2022, December 12). CBER Surveillance Program Biologics Effectiveness and Safety Initiative, Retrieved from https://bestinitiative.org/wp-content/uploads/2022/12/C19-Active-Monitoring-Protocol-Addendum-2022.pdf.

11. Kulldorff, M., et al., A maximized sequential probability ratio test for drug and vaccine safety surveillance. Sequential analysis, 2011. 30(1): p. 58–78

12. Center for Biologics Evaluation and Research, BEST Initiative (2021, Jan 12). Background Rates of Adverse Events of Special Interest for COVID-19 Vaccine Safety Monitoring, Retrieved from https://www.bestinitiative.org/wp-content/uploads/2022/01/C19-Vax-Safety-AESI-Bkgd-Rate-Protocol-FINAL-2020.pdf

13. 13. Wong, H.-L., et al. (2023) Surveillance of covid-19 vaccine safety among elderly persons aged 65 years and older, Vaccine. Available at: https://www.ncbi.nlm.nih.gov/pmc/articles/PMC9712075/ (Accessed: 08 June 2023).

14. Sexson Tejtel, S. K., Munoz, F. M., Al-Ammouri, I., Savorgnan, F., Guggilla, R. K., Khuri-Bulos, N., Phillips, L., & Engler, R. (2022). Myocarditis and pericarditis: Case definition and guidelines for data collection, analysis, and presentation of immunization safety data. Vaccine, 40(10), 1499–1511. 10.1016/j.vaccine.2021.11.074

15. Witberg G, Barda N, Hoss S, et al. Myocarditis after Covid-19 vaccination in a large health care organization. N Engl J Med. 2021;385(23):2132–2139

16. Gargano JW, Wallace M, Hadler SC, et al. Use of mRNA COVID-19 vaccine after reports of myocarditis among vaccine recipients: update from the Advisory Committee on Immunization Practices - United States, June 2021. MMWR Morb Mortal Wkly Rep. 2021;70(27):977–982

17. Nygaard U, Holm M, Bohnstedt C, et al. Population-based incidence of myopericarditis after COVID-19 vaccination in Danish adolescents. Pediatr Infect Dis J. 2022;41(1):e25–e28

18. Centers for Disease Control and Prevention. (2022, June 3). Clinical considerations: Myocarditis and Pericarditis after Receipt of mRNA COVID-19 vaccines among Adolescents and Young Adults. Centers for Disease Control and Prevention. Retrieved August 12, 2022, from https://www.cdc.gov/vaccines/covid-19/clinical-considerations/myocarditis.html

19. Shimabukuro, T. T. (2022). Update on myocarditis following mRNA COVID-19 vaccination.

20. Hause, A. M., Baggs, J., Marquez, P., Myers, T. R., Gee, J., Su, J. R., … & Shay, D. K. (2021). COVID-19 vaccine safety in children aged 5–11 years—United States, November 3–December 19, 2021. Morbidity and Mortality Weekly Report, 70(51-52), 1755.

21. Woodworth, K. R., Moulia, D., Collins, J. P., Hadler, S. C., Jones, J. M., Reddy, S. C., … & Oliver, S. E. (2021). The advisory committee on immunization practices’ interim recommendation for use of Pfizer-BioNTech COVID-19 vaccine in children aged 5–11 years—United States, November 2021. Morbidity and Mortality Weekly Report, 70(45), 2020.

22. Hause AM, Marquez P, Zhang B, et al. COVID-19 mRNA Vaccine Safety Among Children Aged 6 Months–5 Years — United States, June 18, 2022–August 21, 2022. MMWR Morb Mortal Wkly Rep 2022; 71:1115–1120. DOI: 10.15585/mmwr.mm7135a3.

23. United States Food and Drug Administration. Vaccines and Related Biological Products Advisory Committee Meeting June 15, 2022, FDA Briefing Document: EUA amendment request for Pfizer-BioNTech COVID-19 Vaccine for use in children 6 months through 4 years of age. 2022. (Accessed 7 December 2022). https://www.fda.gov/media/159195/download

24. Wang Z, Fang X, et al. Safety and Tolerability of COVID-19 Vaccine in Children With Epilepsy: A Prospective, Multicenter Study. Pediatr Neurol. 2023 Mar;140:3–8. doi: 10.1016/j.pediatrneurol.2022.11.018 Epub 2022 Dec 5. PMID: 36577181; PMCID: PMC9721163.

25. Yang X, Wu L, et al. COVID-19 vaccination for patients with benign childhood epilepsy with centrotemporal spikes. Epilepsy Behav. 2022 Sep;134:108744. doi: 10.1016/j.yebeh.2022.108744 Epub 2022 May 17. PMID: 35952506; PMCID: PMC9110311.

26. Kawai, A. T., Martin, D., Henrickson, S. E., Goff, A., Reidy, M., Santiago, D., Selvam, N., Selvan, M., McMahill-Walraven, C., & Lee, G. M. (2019). Validation of febrile seizures identified in the Sentinel Post-Licensure Rapid Immunization Safety Monitoring Program. Vaccine, 37(30), 4172– 4176. 10.1016/j.vaccine.2019.05.042

27. Centers for Disease Control and Prevention. (2022, October 14). CDC reports early increases in seasonal flu activity. Centers for Disease Control and Prevention. https://www.cdc.gov/flu/spotlights/2022-2023/early-flu-activity.htm#:~:text=October%2014%2C%202022%E2%80%94CDC's%20first,parts%20of%20the%20United%20States.

28. Tang J, et al. Relationship between common viral upper respiratory tract infections and febrile seizures in children from Suzhou, China. J Child Neurol. 2014

29. 29. Van Zeijl, J. H., Mullaart, R. A., Borm, G. F., & Galama, J. M. (2004). Recurrence of febrile seizures in the respiratory season is associated with influenza A. The Journal of pediatrics, 145(6), 800–805

30. Espinoza JA, et al. Impaired learning resulting from respiratory syncytial virus infection. Proc Natl Acad Sci U S A. 2013

31. Sapkota, S., Caruso, E., Kobau, R., Radhakrishnan, L., Jobst, B., DeVies, J., Tian, N., Hogan, R. E., Zack, M. M., & Pastula, D. M. (2022). Seizure- or Epilepsy-Related Emergency Department Visits Before and During the COVID-19 Pandemic - United States, 2019-2021. MMWR. Morbidity and mortality weekly report, 71(21), 703–708. 10.15585/mmwr.mm7121a2

