## Supplemental Tables and Figure for "Safety of Monovalent BNT162b2 (Pfizer-BioNTech), mRNA-1273 (Moderna), and NVX-CoV2373 (Novavax) COVID-19 Vaccines in US Children Aged 6 months to 17 years"

Supplement (Online-only material)

**Supplemental Table 1**. Description of Database Characteristics

**Supplemental Table 2**. Monovalent COVID-19 Vaccines Dosing Schedule Associated with Emergency Use Authorizations or Approval

Description of Database Characteristics

**Supplemental Table 3**. Outcomes, Age Groups, Settings, Clean Windows, Risk Windows, and Analysis Type for the BNT162b2, mRNA-1273, and NVX-CoV2373 COVID-19 Vaccinated Population (Ages 6 months to 17 years)

**Supplemental Table 4**. Administrative Codes for COVID-19 Vaccine Administration used in claims and IIS databases

**Supplemental Table 5.** Descriptive Summary of Outcomes Included in Descriptive Analysis Only

**Supplemental Table 6.** Annual Rates of Seizures/Convulsions Per 100,000 Person-Years by Data Source for Children Aged 2-4 Years and 2-5 Years, 2020 and 2022

**Supplemental Figure 1.** Distribution of Days Between Vaccination and Seizures/Convulsions (N =72 Seizures/Convulsions Events) After Monovalent BNT162b2 (2-4 years) and mRNA-1273 (Ages 2-5 years) COVID-19 Vaccines, Total for All Data Sources

### **Supplemental Table 1. Description of Database Characteristics**

| **Database** | **Description** | **Claims Type** | **Update frequency** | **Data Lag, Time to 80% Completeness*** | **Average No. Enrollees aged 6 months-17 years (2020-2023) ^#^** | **No. IIS Jurisdictions Incorporated into Analysis** |
| --- | --- | --- | --- | --- | --- | --- |
| CVS Health | CVS Health transforms Aetna health plan enrollment, demographic, and medical and drug claims data, for individuals enrolled from January 2018 forward. The Aetna data includes commercial, including Affordable Care act (ACA) Marketplace, and Medicare Advantage health plans into a patient-centered, comprehensive Common Data Model (CDM). | Fully Adjudicated | Monthly | ~ 3-4 months for IP claims, 2-3 months for OP claims, and 1-2 months for professional claims | 6 months-4 years: > 1.08m 5-11 years: > 1.47m 12-15 years: > 934k 16-17 years: > 514k | 23 |
| Optum Pre-adjudicated Claims | The Optum data includes enrollment, prescription drug and pre-adjudicated hospital and physician health insurance claims. The pre-adjudicated claims database includes claims for privately insured and Medicare Advantage enrollees. Hospital and physician claims undergo initial processing on a daily basis from a large number of providers across the US who accept patients with health insurance. | Pre-Adjudicated | Bi-Weekly | ~ 1-2 months for IP, OP, and professional claims | 6 months-4 years: > 839k 5-11 years: > 1.18m 12-15 years: > 750k 16-17 years: > 392k | 20 |
| Carelon Research | Carelon Research is a wholly-owned, research subsidiary of Elevance Health, Inc., a holding company owning several large US health plans associated with Anthem Blue Cross Blue Shield. Carelon has a US population database including individually insured by commercial and Medicare Advantage plans, the Healthcare Integrated Research Database (HIRD), with longitudinal data on health plan enrollees. | Fully Adjudicated | Monthly | ~ 2-3 months for IP claims and 1-2 months for OP and professional claims | 6 months-4 years: > 1.32m 5-11 years: > 1.86m 12-15 years: > 1.20m 16-17 years: > 647k | 9 |

**^*^** Data lag based on 2020 claims delay distribution

^#^Average number of annual enrollees in a given age category is reported for the years 2020-2023 except for Optum where the average is reported through 2021 based on the available data.

### **Supplemental** Table 2. Monovalent COVID-19 Vaccines Dosing Schedule Associated with Emergency Use Authorizations or Approval

| **Brand** | **Age Group (years)** | **Authorized dose** | **Date of Authorization/Approval** |
| --- | --- | --- | --- |
| BNT162b2 | 6 months – 4 | Three-dose primary series (3 micrograms) | EUA: 06/17/2022^[1]^ |
|  | 5-11 | Two-dose primary series (10 micrograms) | EUA (primary series): 10/29/2021^[2]^ |
|  |  | Third primary series dose for immunocompromised individuals (10 micrograms) | EUA (third dose): 01/03/2022^[2]^ |
|  |  | Single booster at least 5 months after completion of primary series (10 micrograms) | EUA (single booster): 05/17/2022 ^[3]^ |
|  | 12-15 | Two-dose primary series (30 micrograms) | EUA (primary series): 05/10/2021^[4]^ |
|  |  | Third primary series dose for immunocompromised individuals (30 micrograms) | EUA (third dose): 08/12/2021^[5]^ |
|  |  | First booster at least 5 months after completion of primary series (30 micrograms) | EUA (first booster): 01/03/2022^[6]^ |
|  |  | Second booster at least 4 months after receipt of first booster for immunocompromised individuals (30 micrograms) | EUA (second booster): 03/29/2022^[6]^ |
|  | 16-17 | Two-dose primary series (30 micrograms) | Approval (primary series): 08/23/2021^[7]^  EUA (primary series): 12/11/2020 ^[7]^ |
|  |  | Third primary series dose for immunocompromised individuals (30 micrograms) | EUA (third dose): 08/12/2021^[5]^ |
|  |  | First booster at least 5 months after completion of primary series (30 micrograms) | EUA (first booster): 12/9/2021^[6]^ |
|  |  | Second booster at least 4 months after receipt of first booster for immunocompromised individuals (30 micrograms) | EUA (second booster): 03/29/2022^[6]^ |
| mRNA-1273 | 6 months – 5 | Two-dose primary series (25 micrograms) | EUA (primary series): 06/17/2022^[8]^ |
|  |  | Third primary series dose for immunocompromised individuals (25 micrograms) | EUA (third dose): 06/17/2022^[8]^ |
|  | 6-11 | Two-dose primary series (50 micrograms) | EUA (primary series): 06/17/2022^[8]^ |
|  |  | Third primary series dose for immunocompromised individuals (50 micrograms) | EUA (third dose): 06/17/2022^[8]^ |
|  | 12-15 | Two-dose primary series (100 micrograms) | EUA (primary series): 06/17/2022^[8]^ |
|  |  | Third primary series dose for immunocompromised individuals (100 micrograms) | EUA (third dose): 06/17/2022^[8]^ |
|  | 16-17 | Two-dose primary series (100 micrograms) | EUA (primary series): 06/17/2022^[8]^ |
|  |  | Third primary series dose for immunocompromised individuals (100 micrograms) | EUA (third dose): 06/17/2022^[8]^ |
| NVX-CoV2373 | 12-17 | Two dose primary series (5 micrograms) | EUA (primary series):  08/19/2022^[9]^ |

### **Supplemental Table 3. Outcomes, Settings, Clean Windows, Risk Windows, and Analysis Type for the BNT162b2, mRNA-1273, and NVX-CoV2373 COVID-19 Vaccinated Population (Ages 6 months to 17 years)**

| **Outcomes*** | **Care Setting** | **Clean Window**** | **Risk Window** | **Analysis Type***** |
| --- | --- | --- | --- | --- |
| Acute Myocardial Infarction | IP | 365 days | 1-28 days ^[10,11]^ | Descriptive Only |
| Anaphylaxis | IP, OP-ED | 30 days | 0-1 day ^[12,13]^ | Descriptive and Sequential Testing |
| Appendicitis | IP, OP-ED | 365 days | 1-42 days ^[14,15]^ | Descriptive and Sequential Testing |
| Bell’s Palsy | IP, OP, PB | 183 days | 1-42 days ^[16]^ | Descriptive and Sequential Testing |
| Common Site Thrombosis with Thrombocytopenia | [Defined in Footnote] ^†^ | 365 days | 1-28 days ^[17]^ | Descriptive and Sequential Testing |
| Deep Vein Thrombosis | IP, OP, PB | 365 days | 1-28 days ^[18,19]^ | Descriptive and Sequential Testing |
| Disseminated Intravascular Coagulation | IP, OP-ED | 365 days | 1-28 days ^[20]^ | Descriptive and Sequential Testing |
| Encephalitis or Encephalomyelitis | IP | 183 days | 1-42 days ^[21]^ | Descriptive and Sequential Testing |
| Febrile Seizures | IP, OP PB | 42 days | 0-1 days** | Descriptive Only |
| Guillain-Barré Syndrome | IP – Primary Position Only | 365 days | 1-42 days ^[22,23]^ | Descriptive and Sequential Testing^#^ |
| Hemorrhagic Stroke | IP | 365 days | 1-28 days ^[10,11]^ | Descriptive and Sequential Testing^#^ |
| Immune Thrombocytopenia | IP, OP, PB | 365 days | 1-42 days ^[24,25]^ | Descriptive and Sequential Testing |
| Kawasaki Disease | IP, OP, PB | 365 days | 1-28 days** | Descriptive Only |
| Multisystem Inflammatory Syndrome in Children | IP, OP-ED | 365 days | 1-42 days ^[26]^ | Descriptive Only |
| Myocarditis or Pericarditis (All Settings) (1-7 day) | IP, OP, PB | 365 days | 1-7 days ^[27]^ | Descriptive and Sequential Testing |
| Myocarditis or Pericarditis (All Settings) (1-21 day) | IP, OP, PB | 365 days | 1-21 days ^[28]^ | Descriptive and Sequential Testing |
| Myocarditis or Pericarditis (IP/OP-ED) (1-7 day) | IP, OP-ED | 365 days | 1-7 days ^[27]^ | Descriptive and Sequential Testing |
| Myocarditis or Pericarditis (IP/OP-ED) (1-21 day) | IP, OP-ED | 365 days | 1-21 days ^[28]^ | Descriptive and Sequential Testing |
| Narcolepsy | IP, OP, PB | 365 days | 1-42 days ^[29-31]^ | Descriptive and Sequential Testing^#^ |
| Non-Hemorrhagic Stroke | IP | 365 days | 1-28 days ^[10, 11]^ | Descriptive and Sequential Testing |
| Pulmonary Embolism | IP, OP, PB | 365 days | 1-28 days ^[18,19,32-35]^ | Descriptive and Sequential Testing |
| Seizures/Convulsions | IP, OP-ED | 42 days | 0-7 days ^[33]^ | Descriptive and Sequential Testing |
| Transverse Myelitis | IP, OP-ED | 365 days | 1-42 days ^[34]^ | Descriptive Only |
| Unusual Site Thrombosis with Thrombocytopenia | [Defined in Footnote] ^†^ | 365 days | 1-28 days ^[35]^ | Descriptive Only |

**Definitions**:

- **Clean Window** is the interval used to define incident outcomes where an individual enters the study cohort only if the health outcome of interest did not occur during that interval.
- **Risk Window** is the interval during which occurrence of the health outcome of interest will be included in the analyses.
- **Care Setting:** IP refers to inpatient facility claims. OP-ED refers to a subset of outpatient facility claims occurring in the emergency department. OP/PB refers to all outpatient facility claims, and professional/provider claims except those professional/provider claims with a laboratory place of service

* Source of the claims-based algorithm: FDA. Appendix 7. Safety AESI Codes. <https://www.bestinitiative.org/wp-content/uploads/2022/01/C19-Vax-Safety-AESI-Bkgd-Rate-Est-Protocol-Suppl.xlsx>

** References for the duration of these windows could not be located in the literature and are instead based on input from clinicians.

*** Sequential testing was conducted only when the minimum threshold of observed events was met (at least 3 events). For NVX-CoV2373 (ages 12 to 17 years) and mRNA-1273 (ages 6 to 17 years) sequential testing did not initiate due to not meeting this minimum threshold for any of the outcomes specified above

^†^ Both Common thrombosis with thrombocytopenia and Unusual site thrombosis (broad) with thrombocytopenia are combined outcomes consisting of a thrombotic event (made up of other events such as acute myocardial infarction, deep vein thrombosis etc.,) and a thrombocytopenia event (defined in the IP, OP/PB setting). The overall setting

definition for each outcome depends on individual setting definitions for each of these components.

^#^ Feasibility of sequential testing of certain outcomes in specific age groups was evaluated based on availability of background rates. Guillain-Barré Syndrome and Hemorrhagic Stroke were sequentially tested in ages <5 years only and Narcolepsy in ages >5 years old only due to lack of estimable background rates in other age groups

### **Supplemental** Table 4. Administrative Codes for COVID-19 Vaccine Administration used in claims and IIS databases

| **HCPCS/CPT* Codes** | **CVX** Codes (IIS-Specific)** | **Manufacturer** | **Name** | **Age Group (years)** | **Vaccine Administration Code** | **NDC*** 11 Labeler Product ID (Vial)** | **Dosing Interval** |
| --- | --- | --- | --- | --- | --- | --- | --- |
| 91308 | 219 | Pfizer | Pfizer-BioNTech COVID-19 Vaccine | 6 m- 4 | 0081A (1st dose) | 59267-0078-01 59267-0078-04 | - 21+ days between dose 1 and dose 2 - 28+ days between dose 2 and dose 3 |
|  |  |  |  |  | 0082A (second dose) |  |  |
|  |  |  |  |  | 0083A (third dose) |  |  |
| 91307 | 218 | Pfizer | Pfizer-BioNTech COVID-19 Pediatric Vaccine | 5-11 | 0071A (1st dose) | 59267-1055-0159267-1055-02 59267-1055-04 | - 21+ days between dose 1 and dose 2 - For immunocompromised, 21+ days between dose 1 and dose 2, and 28+ days between dose 2 and additional primary dose (dose 3) |
|  |  |  |  |  | 0072A (2nd dose) |  |  |
|  |  |  |  |  | 0073A (3rd dose) |  |  |
|  |  |  |  |  | 0074A (booster dose) |  |  |
| 91305 | 217 | Pfizer | Pfizer-BioNTech COVID-19 Vaccine | 12-17 | 0051A (1^st^ dose) | 59267-1025-01 59267-1025-02 59267-1025-03 59267-1025-04 | - 21+ days between dose 1 and dose 2 and 5+ months between dose 2 and third/booster dose - For immunocompromised, 21+ days between dose 1 and dose 2, 28+ days between dose 2 and dose 3. Additionally, booster dose recommended 3+ months after primary series |
|  |  |  |  |  | 0052A (2^nd^ dose) |  |  |
|  |  |  |  |  | 0053A (3^rd^ dose) |  |  |
|  |  |  |  |  | 0054A (booster dose) |  |  |
| 91300 | 208 | Pfizer | Pfizer-BioNTech COVID-19 Vaccine | 12-17 | 0001A (1^st^ dose) | 59267-1000-01 59267-1000-02 59267-1000-03 | - 21+ days between dose 1 and dose 2 and 5+ months between dose 2 and third/booster dose - For immunocompromised, 21+ days between dose 1 and dose 2, 28+ days between dose 2 and dose 3. Additionally, booster dose recommended 3+ months after primary series |
|  |  |  |  |  | 0002A (2^nd^ dose) |  |  |
|  |  |  |  |  | 0003A (3^rd^ dose) |  |  |
|  |  |  |  |  | 0004A (booster dose) |  |  |
|  |  |  |  |  | 0002A (2^nd^ dose) |  |  |
|  |  |  |  |  | 0003A (3^rd^ dose) |  |  |
|  |  |  |  |  | 0004A (booster dose) |  |  |
| 91301, 91306, 91309 | 207 | Moderna | Moderna COVID-19 Vaccine | 6 m-17 | 0011A, 0091A (1^st^ dose) | 61434-0043-00  61434-0043-01  80777-0100-11  80777-0100-98  80777-0100-99  80777-0277-05  80777-0279-05  80777-0273-98  80777-0273-10  80777-0273-99  80777-0273-15  80777-0273-98  80777-0273-05  80777-0273-99 | - 28+ days between dose 1 and dose 2 - For immunocompromised, 28 days between dose 1 and dose 2, and 28+ days between dose 2 and dose 3. |
|  |  |  |  |  | 0012A, 0092A  (2^nd^ dose) |  |  |
|  |  |  |  |  | 0013A, 0093A  (3^rd^ dose) |  |  |
|  |  |  |  |  | 0064A, 0094A (booster dose) |  |  |
| 91304 | 211 | Novavax | Novavax COVID-19 Vaccine | 12-17 | 0041A (1^st^ dose) | 80631-0100-01  80631-0100-10 | - 21+ days between dose 1 and dose 2 including for immunocompromised |
|  |  |  |  |  | 0042A  (2^nd^ dose) |  |  |

* HCPCS: The Healthcare Common Procedure Coding System produced by the Centers for Medicare and Medicaid Services (CMS)^[36]^

CPT: The Current Procedural Terminology codes^[36]^

** CVX: Vaccine administered code set developed and maintained by the Centers for Disease Control and Prevention’s (CDC) National Center of Immunization and Respiratory Diseases (NCIRD)^[37]^

*** NDC: National Drug Codes used to identify and report drugs using a unique, three-segment number which serves as the Food and Drug Administration’s (FDA) identifier for drugs^[38]^

### **Supplemental Table 5. Descriptive Summary of Outcomes Included in Descriptive Analysis Only**

| **Outcome** | **Brand** | **All Data Sources** | | **Carelon Research^a^** | | **CVS Health^b^** | | **Optum^c^** | |
| --- | --- | --- | --- | --- | --- | --- | --- | --- | --- |
|  |  | **No. Vaccine Doses** | **No. Outcomes** | **No. Vaccine Doses** | **No. Outcomes** | **No. Vaccine Doses** | **No. Outcomes** | **No. Vaccine Doses** | **No. Outcomes** |
| Acute myocardial infarction | BNT162b2 | 6,650,342 | § | 2,589,974 | § | 2,120,805 | § | 1,939,563 | 0 |
|  | mRNA-1273 | 252,959 | 0 | 94,878 | 0 | 85,473 | 0 | 72,608 | 0 |
|  | NVX-CoV2373 | 98 | 0 | 18 | 0 | 18 | 0 | 62 | 0 |
| Febrile Seizure | BNT162b2 | 7,932,684 | 76 | 3,064,741 | 26 | 2,565,776 | 27 | 2,302,167 | 23 |
|  | mRNA-1273 | 315,520 | 37 | 119,174 | 14 | 108,685 | § | 87,661 | § |
|  | NVX-CoV2373 | 133 | 0 | 21 | 0 | 28 | 0 | 84 | 0 |
| Guillain-Barré syndrome | BNT162b2 | 6,650,324 | § | 2,589,971 | § | 2,120,793 | § | 1,939,560 | § |
|  | mRNA-1273 | 252,959 | 0 | 94,878 | 0 | 85,473 | 0 | 72,608 | 0 |
|  | NVX-CoV2373 | 98 | 0 | 18 | 0 | 18 | 0 | 62 | 0 |
| Hemorrhagic stroke | BNT162b2 | 6,650,157 | 14 | 2,589,902 | § | 2,120,744 | § | 1,939,511 | § |
|  | mRNA-1273 | 252,950 | 0 | 94,874 | 0 | 85,473 | 0 | 72,603 | 0 |
|  | NVX-CoV2373 | 98 | 0 | 18 | 0 | 18 | 0 | 62 | 0 |
| Kawasaki disease | BNT162b2 | 6,648,712 | 79 | 2,589,367 | 26 | 2,120,267 | 23 | 1,939,078 | 30 |
|  | mRNA-1273 | 252,810 | § | 94,809 | § | 85,441 | § | 72,560 | § |
|  | NVX-CoV2373 | 98 | 0 | 18 | 0 | 18 | 0 | 62 | 0 |
| Multisystem inflammatory syndrome in children (MIS-C) | BNT162b2 | 6,650,003 | 15 | 2,589,829 | § | 2,120,710 | § | 1,939,464 | § |
|  | mRNA-1273 | 252,949 | 0 | 94,878 | 0 | 85,469 | 0 | 72,602 | 0 |
|  | NVX-CoV2373 | 98 | 0 | 18 | 0 | 18 | 0 | 62 | 0 |
| Narcolepsy | BNT162b2 | 6,649,400 | 65 | 2,589,563 | 26 | 2,120,525 | 22 | 1,939,312 | 17 |
|  | mRNA-1273 | 252,954 | 0 | 94,877 | 0 | 85,472 | 0 | 72,605 | 0 |
|  | NVX-CoV2373 | 98 | 0 | 18 | 0 | 18 | 0 | 62 | 0 |
| Transverse myelitis | BNT162b2 | 6,650,321 | § | 2,589,967 | § | 2,120,801 | 0 | 1,939,553 | § |
|  | mRNA-1273 | 252,959 | § | 94,878 | § | 85,473 | 0 | 72,608 | 0 |
|  | NVX-CoV2373 | 98 | 0 | 18 | 0 | 18 | 0 | 62 | 0 |
| Unusual site thrombosis with thrombocytopenia | BNT162b2 | 6,650,303 | § | 2,589,961 | § | 2,120,793 | § | 1,939,549 | 0 |
|  | mRNA-1273 | 252,959 | 0 | 94,878 | 0 | 85,473 | 0 | 72,608 | 0 |
|  | NVX-CoV2373 | 98 | 0 | 18 | 0 | 18 | 0 | 62 | 0 |

^a^ Data through 3/2023 ^b^ Data through 2/2023 ^c^ Data through 4/2023

^§^ Cell sizes 1-10 and cells that can be used to back-calculate small cell sizes were masked for confidentiality

### Supplemental Table 6. Background Annual Rate of Seizures/Convulsions Per 100,000 Person-Years by Data Source in Children Aged 2-4 Years and 2-5 Years, 2020 and 2022

|  | **Ages 2-4 years** | | | **Ages 2-5 years** | | |
| --- | --- | --- | --- | --- | --- | --- |
| Data Source | **2020** | **2022^a^** | **Difference (Ratio)** | **2020** | **2022^a^** | **Difference (Ratio)** |
| Carelon Research | 234.35 | 551.18 | 2.35 | 207.03 | 470.51 | 2.27 |
| CVS Health | 190.55 | 434.57 | 2.28 | 166.72 | 369.11 | 2.21 |
| Optum | 192.96 | 448.26 | 2.32 | 168.13 | 378.07 | 2.25 |

^a^ For all three data sources, this includes background rates data through 12/2022

### Supplemental Figure 1. Distribution of Days Between Vaccination and Seizures/Convulsions (N =72 Seizures/Convulsions Events) After Monovalent BNT162b2 (2-4 years) and mRNA-1273 (Ages 2-5 years) COVID-19 Vaccines, Total for All Data Sources

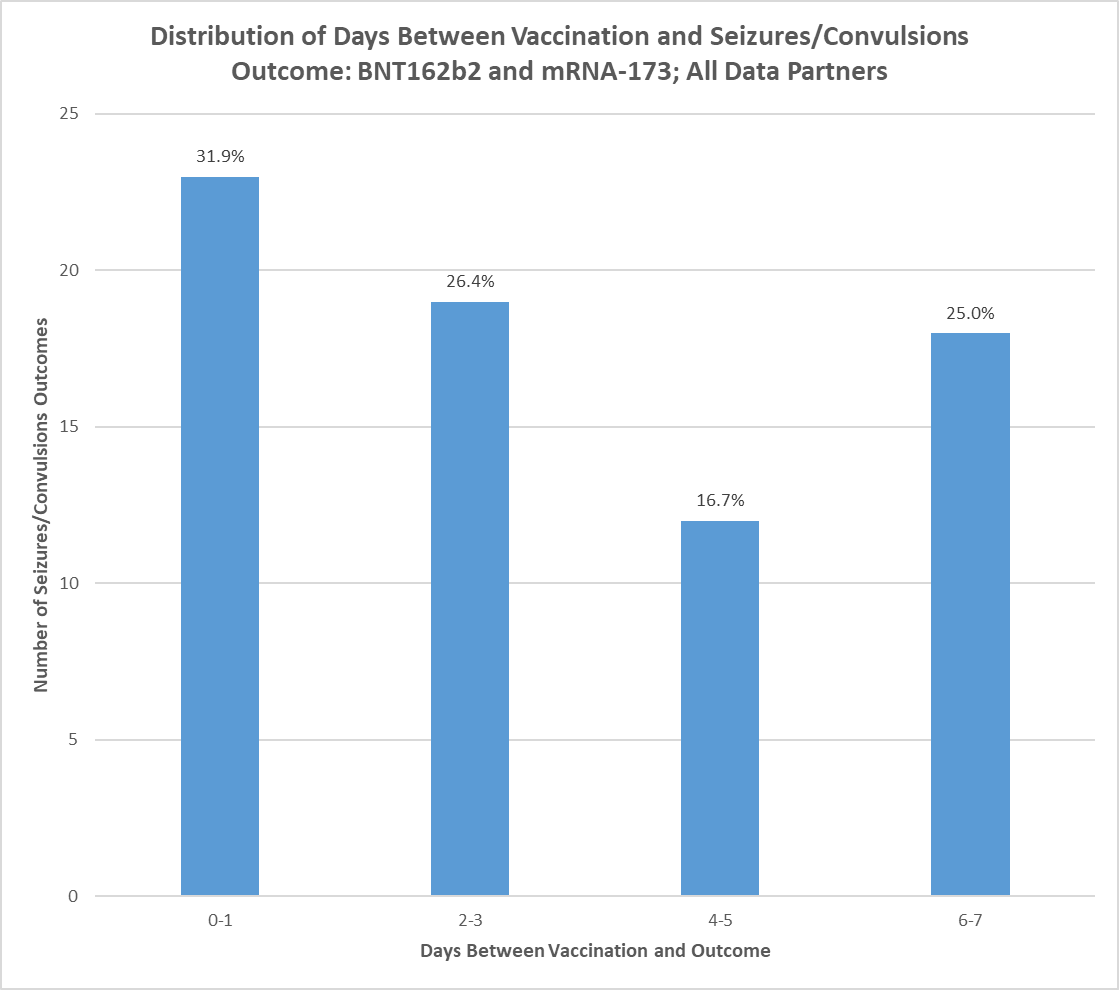

Combines data for all three data sources: Carelon Research: Data through 3/2023, CVS Health: Data through 2/2023, Optum: Data through 4/2023

### References

1. U.S. Food and Drug Administration. (2022, June 16). Pfizer-BioNTech COVID-19 Vaccine EUA Amendment for Use in Individuals 6 Months Through 4 Years of Age. U.S. Food and Drug Administration. Retrieved July 27, 2022, from https://www.fda.gov/media/159393/download
2. U.S. Food and Drug Administration. (2022, January 3). Coronavirus (COVID-19) Update: FDA Takes Multiple Actions to Expand use of Pfizer-BioNtech COVID-19 vaccine. U.S. Food and Drug Administration. Retrieved July 27, 2022, from https://www.fda.gov/news-events/press-announcements/coronavirus-covid-19-update-fda-takes-multiple-actions-expand-use-pfizer-biontech-covid-19-vaccine
3. U.S. Food and Drug Administration. (2022, May 17). Coronavirus (COVID-19) Update: FDA Expands Eligibility for Pfizer-BioNtech COVID-19 Vaccine Booster Dose to Children 5 through 11 Years. U.S. Food and Drug Administration. Retrieved July 27, 2022, from https://www.fda.gov/news-events/press-announcements/coronavirus-covid-19-update-fda-expands-eligibility-pfizer-biontech-covid-19-vaccine-booster-dose
4. U.S. Food and Drug Administration. (2021, May 10). Emergency Use Authorization (EUA) Amendment for an Unapproved Product Review Memorandum. U.S. Food and Drug Administration. Retrieved July 27, 2022, from https://www.fda.gov/media/148542/download
5. Cho, D. (2021, August 12). Review Memorandum. U.S. Food and Drug Administration. Retrieved July 27, 2022, from https://www.fda.gov/media/151613/download
6. O'Shaughnessy, J. A. (2022, July 8). Pfizer-BioNTech COVID-19 Vaccine EUA LOA reissued. U.S. Food and Drug Administration. Retrieved July 27, 2022, from https://www.fda.gov/media/150386/download
7. U.S. Food and Drug Administration. (2021, August 23). FDA Approves First COVID-19 Vaccine. U.S. Food and Drug Administration. Retrieved July 27, 2022, from https://www.fda.gov/news-events/press-announcements/fda-approves-first-covid-19-vaccine
8. O'Shaughnessy, J. A. (2022, June 17). Moderna COVID-19 Vaccine EUA Letter of Authorization. U.S. Food and Drug Administration. Retrieved July 27, 2022, from https://www.fda.gov/media/144636/download
9. U.S. Food and Drug Administration. (2022, August 19). FDA Roundup: August 19, 2022. U.S. Food and Drug Administration. Retrieved December 8, 2022, from https://www.fda.gov/news-events/press-announcements/fda-roundup-august-19-2022
10. Al Qudah Z, Abukwaik W, Souayah N. Stroke after Vaccination in United States. A Report from the CDC/FDA Vaccine Adverse Event Reporting System. [1990–2010] (P01.009). Neurology. 2012; 78(Meeting Abstracts 1): P01.009-P01.009.
11. Smeeth L, Thomas SL, Hall AJ, Hubbard R, Farrington P, Vallance P. Risk of myocardial infarction and stroke after acute infection or vaccination. New England Journal of Medicine. 2004; 351(25): 2611-2618.
12. Rüggeberg JU, Gold MS, Bayas J-M, et al., Anaphylaxis: case definition and guidelines for data collection, analysis, and presentation of immunization safety data. Vaccine. 2007; 25(31): 5675-5684.
13. Su JR, Moro PL, Ng CS, Lewis PW, Said MA, Cano MV. Anaphylaxis after vaccination reported to the Vaccine Adverse Event Reporting System, 1990-2016. Journal of Allergy and Clinical Immunology. 2019; 143(4): 1465-1473.
14. Donahue JG, Kieke BA, Lewis EM, et al. Near Real-Time Surveillance to Assess the Safety of the 9-Valent Human Papillomavirus Vaccine. Pediatrics. 2019; 144(6): e20191808.
15. GeeJ, Naleway A, Shui I et al. Monitoring the safety of quadrivalent human papillomavirus vaccine: findings from the Vaccine Safety Datalink. Vaccine. 2011; 29(46): 8279-84.
16. Renoud L, Khouri C, Revol B, et al. Association of Facial Paralysis With mRNA COVID-19 Vaccines: A Disproportionality Analysis Using the World Health Organization Pharmacovigilance Database. JAMA Internal Medicine. 2021; 181(9): 1243–1245.
17. Pishko AM, Bussel JB, Cines DB. COVID-19 vaccination and immune thrombocytopenia. Nature Medicine. 2021; 27: 1145–1146 .
18. Kearon C. Natural history of venous thromboembolism. Circulation. 2003; 107(23_suppl_1): I-22-I-30.
19. Vickers E.R, McClure DL, Naleway AL, et al. Risk of venous thromboembolism following influenza vaccination in adults aged 50 years and older in the Vaccine Safety Datalink. Vaccine. 2017; 35(43): 5872-5877.
20. Tang N, Li D, Wang X, Sun Z . Abnormal coagulation parameters are associated with poor prognosis in patients with novel coronavirus pneumonia. Journal of Thrombosis Haemostasis. 2020; 18(4): 844-847.
21. Pellegrino P, Carnovale C, Perrone V, et al. Acute disseminated encephalomyelitis onset: evaluation based on vaccine adverse events reporting systems. PloS one. 2013; 8(10): e77766.
22. Dodd CN, Romio SA, Black S, et al. International collaboration to assess the risk of Guillain Barré Syndrome following Influenza A (H1N1) 2009 monovalent vaccines. Vaccine. 2013; 31(40): 4448-4458.
23. Schonberger LB, Bregman DJ, Sullivan-Bolyai JZ, et al. Guillain-Barré syndrome following vaccination in the national influenza immunization program, United States, 1976–1977. American Journal of Epidemiology. 1979; 110(2): 105-123.
24. Black C, Kaye JA, Jick H. MMR vaccine and idiopathic thrombocytopaenic purpura. British Journal of Clinical Pharmacology. 2003; 55(1): 107-111.
25. D'alò GL, Zorzoli E, Capanna A, et al. Frequently asked questions on seven rare adverse events following immunization. Journal of Preventive Medicine Hygiene. 2017; 58(1): E13.
26. Hennon TR, Penque MD, Abdul-Aziz R, et al. COVID-19 associated Multisystem Inflammatory Syndrome in Children (MIS-C) guidelines; a Western New York approach. Progress in Pediatric Cardiology. 2020; 57: 1-6.
27. Oster ME, Shay DK, Su JR, et al. Myocarditis Cases Reported After mRNA-Based COVID-19 Vaccination in the US From December 2020 to August 2021. JAMA. 2022; 327(4): 331–340.
28. Klein N. Myocarditis Analyses in the Vaccine Safety Datalink: Rapid Cycle Analyses and “Head-to-Head” Product Comparisons. Presentation at: Advisory Committee on Immunization Practices; October, 2021.
29. Duffy J, Weintraub E, Vellozzi C, DeStefano F . Narcolepsy and influenza A (H1N1) pandemic 2009 vaccination in the United States. Neurology. 2014; 83(20): 1823-1830.
30. Montplaisir J, Petit D, Quinn M-J, et al. Risk of narcolepsy associated with inactivated adjuvanted (AS03) A/H1N1 (2009) pandemic influenza vaccine in Quebec. PloS one. 2014; 9(9): e108489.
31. Sarkanen TO, Alakuijala APE, Dauvilliers YA, Partinen MM. Incidence of narcolepsy after H1N1 influenza and vaccinations: Systematic review and meta-analysis. Sleep Medicine Reviews. 2018; 38: 177-186.
32. Kearon C, Akl EA. Duration of anticoagulant therapy for deep vein thrombosis and pulmonary embolism. Blood. 2014; 123(12): 1794-1801.
33. Duffy J, Weintraub E, Hambidge SJ, et al. Febrile seizure risk after vaccination in children 6 to 23 months. Pediatrics. 2016; 138(1). e20160320.
34. Agmon-Levin N, Kivity S, Szyper-Kravitz M, Shoenfeld Y., Transverse myelitis and vaccines: a multi-analysis. Lupus. 2009; 18(13): 1198-1204.
35. Whitworth H, Sartain SE, Kumar R, et al. Rate of thrombosis in children and adolescents hospitalized with COVID-19 or MIS-C. Blood. 2021; 138(2): 190-198.
36. License for use of current procedural terminology, Fourth edition (‘Cpt®’) CMS.gov. Available at: https://www.cms.gov/license/ama?file=%2Ffiles%2Fzip%2Flist-codes-effective-january-1-2023-published-december-1-2022.zip (Accessed: 31 August 2023).
37. IIS (2023) Centers for Disease Control and Prevention. Available at: https://www2a.cdc.gov/vaccines/iis/iisstandards/vaccines.asp?rpt=cvx (Accessed: 31 August 2023).
38. (2023) National Drug Code Directory. Available at: https://www.accessdata.fda.gov/scripts/cder/ndc/index.cfm (Accessed: 06 October 2023).
